# Shared and distinct phenotypic profiles among neurodevelopmental disorder genes

**DOI:** 10.64898/2026.02.15.26346328

**Authors:** Hermela Shimelis, Matthew T. Oetjens, Bobbi McGivern, Zhancheng Zhang, Janelle E. Stanton, Ian McSalley, Shiva Ganesan, Brenda M. Finucane, Ingo Helbig, Christa L. Martin, Scott M. Myers, David H. Ledbetter

**Affiliations:** Department of Developmental Medicine, Geisinger, Lewisburg, PA, United States; GeneDx, LLC, Gaithersburg, MD, United States; Florida Institute for Pediatric Rare Diseases, Florida State University College of Medicine, Tallahassee, FL, United States; Division of Neurology, Children’s Hospital of Philadelphia, Philadelphia, PA, United States; Deptartment of Biomedical and Health Informatics (DBHi), Children’s Hospital of Philadelphia, Philadelphia, PA, United States; The Epilepsy NeuroGenetics Initiative (ENGIN), Children’s Hospital of Philadelphia, Philadelphia, PA, United States; Department of Neurology, University of Pennsylvania Perelman School of Medicine, Philadelphia, PA, United States

## Abstract

Rare pathogenic variants in many genes contribute to neurodevelopmental disorders (NDDs), including intellectual disability and/or global developmental delay (ID), autism spectrum disorder (ASD), epilepsy (EP), and cerebral palsy (CP). These conditions frequently co-occur and share genetic etiologies, yet the broader phenotypic effects and the extent of shared versus distinct genetic influences remain unclear. Here, we adopt a cross-disorder framework to examine NDD genes across four diagnostic categories, characterize gene-associated phenotypic profiles, and identify convergent pathways that help refine how pathogenic variants in these genes shapes clinical outcomes. Using a discovery cohort of 8,973 probands with disease-causing variants in 263 NDD genes, we performed phenotype-based gene clustering and identified six distinct gene clusters. These clusters reveal structured patterns of genetic overlap, showing that subsets of NDD genes preferentially contribute to specific disorder combinations of ID, ASD, EP, and CP. The largest gene cluster was characterized by ID, whereas the other five included one enriched for ASD and ID, two for EP and ID and two for CP and ID, each with significantly differing frequencies. In an independent validation cohort of 19,704 probands, five of the six clusters were replicated. Gene Ontology enrichment analyses revealed distinct biological processes in each cluster, suggesting that coherent molecular mechanisms underlie the differing NDD diagnostic profiles. Together these findings demonstrate that NDD genes fall into coherent clusters that consistently map onto characteristic phenotype profiles, providing a framework to inform future therapeutic strategies and support early prognostication for individuals with pathogenic variants in NDD genes.

## Introduction

Neurodevelopmental disorders (NDDs) result from multiple genomic determinants acting in concert with environmental and stochastic developmental factors, producing broad and overlapping clinical phenotypes [1]. Unlike insults to the mature nervous system, which often cause focal neurologic deficits, dysfunction during brain development tends to be diffuse, resulting in varying degrees and patterns of impairment in cognitive, motor, and neurobehavioral functioning and, in some cases, observable morphological brain anomalies or neurophysiological abnormalities [1, 2]. Intellectual disability and/or global developmental delay (ID), autism spectrum disorder (ASD), epilepsy (EP), and cerebral palsy (CP) are prototypical NDDs with predominant impairment in cognitive, neurobehavioral, neurophysiologic, and motor domains, respectively. These conditions are highly heterogeneous and frequently comorbid [1, 2]. Large-scale genomic studies have identified rare deleterious variants in >1,000 genes associated with ID, ASD, CP, and EP [3–12]. However, pathogenic variants in NDD genes rarely confer risk for a single disorder; instead, the same genes are often implicated across multiple NDDs. This heterogeneity and frequent comorbidity complicates efforts to link specific gene disruptions to discrete clinical NDD diagnoses [1, 2, 13, 14].

Efforts to identify genes preferentially associated with specific NDDs, such as ASD-or ID-predominant genes, have largely relied on one-to-one comparisons of *de novo* variants between pairs of clinically ascertained cohorts (ASD versus ID, EP versus ID, etc.) [5, 6, 15, 16]. However, this dichotomization fails to capture the extensive gene overlap and high comorbidity across NDDs [7, 17, 18], leading to incomplete or misleading gene-phenotype associations. Therefore, genes contributing to multiple NDD phenotypes may be misclassified as disorder-specific, obscuring shared mechanisms. Although convergent genetic risk across NDDs suggests common biological pathways, these remain poorly defined [19–22], because each genetic etiology is rare [23]. Systematic evaluation of multiple phenotypes across large gene sets can reveal recurrent biological patterns, improve clinical prognostication—particularly following early genetic diagnosis—and clarify phenotype-specific genetic liabilities that may inform therapeutic development [16, 21, 24–26]. Although such insights would ideally be derived from large, population-based cohorts with comprehensive phenotyping across diagnostic boundaries [17], enabling unbiased assessment of each condition, such resources are not currently available.

Understanding the biological basis of clinical heterogeneity remains a central challenge in NDDs. Phenotype-based clustering has been used to identify subgroups with shared traits [27–29], enrich cohorts for common variants [30, 31], prioritize candidate variants [32], and examine phenotype-genotype comorbidity in NDDs [22, 33, 34]. In genetically defined NDD cohorts, hierarchical clustering of clinical features followed by gene-enrichment analyses has revealed biological processes that differ across subtypes [35]. However, broad phenotypic categories and high variability often produce numerous small clusters, limiting the ability to detect consistent, reproducible patterns. Recent studies have shown that genes implicated in ASD can be clustered based on their biological functions [22, 35, 36], but the exclusion of other NDDs complicates efforts to distinguish ASD-specific mechanisms from those shared with other NDDs. Although initial attempts have been made to include multiple NDD phenotypes (ID, EP, and ASD), capturing comorbid NDDs has remained a conceptual limitation [11].

These limitations underscore the need for integrative approaches that account for comorbidity and enable systematic identification of shared and disorder-specific pathways across NDDs.

Nevertheless, recent large-scale cross-disorder studies of common variants in neurodevelopmental and psychiatric disorders [19, 37–40], as well as other disease areas with substantial genetic overlap [41], have shown that integrative analytical approaches can effectively identify shared and distinct genetic architectures. These cross-disorder genomic studies have been successful in identifying risk alleles that converge on common and distinct biological pathways. In contrast, large-scale cross-disorder analyses of rare pathogenic variants, particularly those capable of defining gene-level phenotypic profiles and distinguishing those that contribute to shared versus distinct NDD trajectories remain limited. This gap persists in part because monogenic NDDs are individually rare, making it difficult to assemble sufficiently powered cohorts needed to discern gene-specific phenotypic profiles across disorders [42].

To address these gaps, we examined patterns of phenotypic convergence across 263 NDD genes in a cohort of 8,973 probands and identified six gene clusters with distinct diagnostic profiles characterized by differing patterns of involvement across cognitive, neurobehavioral, neurophysiologic, and motor domains, reflected by ID, ASD, EP, and CP diagnoses, respectively. Five of the six clusters replicated in an independent cohort, supporting their robustness. Gene Ontology analyses further showed that each cluster is enriched for distinct biological processes. Together, these findings provide a clearer understanding of the molecular mechanism underlying different NDD profiles. They also generate testable hypotheses and underscore the value of large, cross-disorder clinical cohort and phenotype-driven approaches that are grounded in recurring patterns across multiple disorders, rather than single-disorder ascertainment, for elucidating shared and distinct genetic relationships among NDDs. Our findings help advance the field beyond recognizing genetic overlap to defining a structured organization of gene-phenotype relationships and their biological underpinnings.

## Materials and Methods

### Identification of study cohorts and gene sets

The study cohorts are described in Supplementary Materials.

### Phenotype based Hierarchical clustering

We clustered genes based on similarity in phenotype frequency profiles across the four disorders. For each gene, we calculated the percentage of individuals diagnosed with ID, ASD, EP and CP. These gene-level phenotype frequencies were used as input for hierarchical agglomerative clustering. Prior to clustering, data were pre-processed by applying standard scaling and Principal Component Analysis (PCA), retaining all four principal components.

### Optimizing distance metrics and linkage methods

To select the optimal distance metric and linkage method for hierarchical clustering, we tested multiple combinations and chose the one with the highest Cophenetic correlation coefficient, which was achieved using the Manhattan distance and average linkage method. The optimal linkage-distance threshold was determined by testing a range of values; a threshold of 4.0 was selected based on optimal performance across two cluster quality metrics: Silhouette Score, and Calinski-Harabasz index. Flat clusters were then generated using SciPy’s fcluster function with this distance threshold, yielding seven clusters, including one single gene cluster. Each gene was assigned to one cluster, and clusters were labeled according to phenotype(s) with a median frequency;= 25% across genes within the cluster

### Assessment of cluster stability

Cluster stability was evaluated using bootstrap resampling. Hierarchical clustering was repeated 50 times on random subsets containing 50% or 80% of the 263 genes (sampled with replacement), using Manhattan distance and average linkage. For each bootstrap, the optimal partition was selected by maximizing the Calinski-Harabasz index across distance thresholds. Pairwise similarity between bootstrap cluster solutions was quantified with the Adjusted Rand Index (ARI), and overall clustering stability was summarized by the mean and standard deviation of ARI values. The 50% subsample produced a mean ARI of 0.93 ± 0.26, and the 80% subsample produced a mean ARI of 0.99. To assess cluster stability, we also conducted a permutation test with 1,000 random label shuffling. For each permutation, cluster labels were permuted without replacement and the ARI between the permuted labels and the original cluster labels was computed to estimate similarity expected by chance. All clusters yielded p-value < 0.0001, statistically robust clustering beyond random chance.

### Statistical evaluation of phenotype profile differences across gene clusters

Gene cluster phenotype profiles were defined as the median frequency of each of the four disorders across the genes within the cluster. The 95% confidence intervals (CI) for median frequencies were estimated using bootstrap resampling with replacement (10,000 iterations). To test whether disorder frequencies differed from the overall gene set, we applied two-sided one-sample Wilcoxon signed-rank tests; between cluster differences were assessed using pairwise Mann-Whitney U tests. P-values were corrected for multiple testing across all cluster-phenotype comparisons using the Benjamini-Hochberg procedure (FDR = 5%).

### Validation of gene cluster in an independent cohort

We applied a result-based cluster validation approach, as previously described [43, 44], to assign 234 genes shared between the discovery and validation cohorts to the six phenotype-defined clusters.

Gene clusters were originally derived in the discovery dataset using hierarchical clustering, and the single-gene cluster (*DEAF1)* was excluded from the validation analyses. For validation, gene-level phenotype frequencies (ID, ASD, EP, and CP) were obtained from the validation cohort and preprocessed using the same PCA-based transformation applied to the discovery data. Five supervised classifiers (K-nearest Neighbors, Decision Tree, Random Forest, Support Vector Machine (SVM), Logistic Regression) were trained on the discovery dataset to predict cluster membership in the validation dataset. Hyperparameters were optimized using 5-fold cross-validation, and the Random Forest model, which comprised 200 trees with default splitting parameters was selected based on performance (mean accuracy of 0.954, SD = 0.031). The optimized model was then applied to the validation dataset to predict cluster assignments for 234 genes. Agreement between predicted and discovery labels was quantified using accuracy and ARI. Confusion matrix was constructed to visualize concordance between discovery and validation cluster labels. Phenotype profiles for each predicted cluster were generated by summarizing phenotype frequencies and gene counts in the validation dataset. All analyses were performed in Python version 3.12.2.

### Sensitivity analysis using a subset of the validation cohort

We evaluated potential overlap between the discovery and validation cohorts, as the discovery cohort was assembled from published studies, some of which included GeneDx NDD cases. Seven such studies were identified, accounting for 475 of 19,704 probands (2.4%) in the validation cohort.

Because individual-level identifiers were unavailable, these cases could not be excluded from the validation cohort. To mitigate any impact, we repeated the validation analyses using a non-overlapping time window. For sensitivity analysis, we repeated cluster validation using a non-overlapping subset of 4,192 individuals with variants in 149 genes, ensuring no shared cases between the discovery and validation cohorts.

### Statistical tests for evaluating whether cluster predictions exceeded chance

We used permutation test to assess whether agreement between predicted validation and discovery cluster exceeded chance. First, we calculated the observed number of matching labels between the two vectors. To generate a null distribution, predicted labels were permuted 10,000 times while keeping discovery labels fixed, and matches were recalculated. The empirical p-value was the proportion of permutations with matches greater than or equal to the observed count. To estimate agreement expected by chance, we computed the expected number of matching labels under random assignment. For each cluster label, we multiplied its frequency in the discovery and predicted (validation) label vectors and divided by the sample size (N = 234). Summing these values across labels yielded an expected 75.2 random matches, compared with 155 matches observed.

To test agreement within each cluster, we performed cluster-wise permutation test. For each cluster, we counted the number with matching labels (observed matches). We then permuted the full vector of predicted labels 10,000 times, preserving overall label frequencies, and recalculated matches within that cluster for each permutation. One-sided empirical p-values were computed as the proportion of permutations with matches greater than or equal to the observed value. Expected matches were defined as the mean across permutations.

### Analysis of functional enrichment within gene clusters

Gene ontology (GO) Biological Processes (BPs) enrichment analysis was performed for each gene cluster using the clusterProfiler [45, 46] package in R. Only GO terms shared by at least two genes within a cluster were included. Redundant GO terms were removed with the Simplify function (similarity cutoff = 0.7; select_fun = “miin”). The top enriched terms were visualized using dot plot showing the ten GO BPs with the lowest Benjamini & Hochberg (“BH”) adjusted p-values. Fold enrichment was calculated relative to all protein-coding genes as the background Gene-Concept networks were generated using cnetplot to visualize relationship between genes and significantly enriched GO terms [47]. Functional similarity between clusters was assessed with GOSemSim, using each cluster’s enriched GO terms [48, 49]. Pairwise semantic similarity scores were computed with mgoSim (Wang method, Best-Match Average) to quantify functional relatedness among clusters.

### Reactome pathway enrichment analysis

REACTOME pathway enrichment was performed using ReactomePA R package [50]. Only pathway terms shared by at least two genes in a cluster were included. The top ten enriched pathways were visualized using dot plots showing the lowest Benjamini & Hochberg (“BH”) adjusted p-values. Gene-Concept networks were generated to visualize relationship between genes and enriched pathways.

## Results

### Identifying genetic etiologies with similar phenotypic profiles

To assess whether NDD genes organize into groups defined by shared phenotypic profiles, we analyzed a large discovery cohort comprising 8,973 NDD probands diagnosed with at least one of four disorders (ID, ASD, EP, or CP) and harboring a disease-causing variant in one of 263 high-confidence genes (Supplementary Tables 1 and 2; Supplementary Fig.1). Hypothesis-free hierarchical clustering of gene-level phenotype frequencies revealed six distinct gene clusters: ID, ASD–ID, ID–EP, EP–ID, ID–CP, and CP (Fig.1; Fig. 2a). One gene (*DEAF1*) did not cluster and was excluded from downstream analyses. Clusters were named based on phenotypes with median frequencies of >25% across genes within the cluster, ordered from highest to lowest (e.g., ASD–ID cluster). The hierarchical dendrogram shows how genes segregate into the six clusters based on similarity in their phenotype frequency profiles, suggesting the presence of coherent phenotype-aligned groups. Cluster structure was supported by optimal clustering metrics (Silhouette score = 0.307; Calinski-Harabasz index = 104.0; Supplementary Fig. 2). Two-dimensional projection of gene-phenotype matrix provides a visual representation of separation and relationships among clusters identified by hierarchical clustering, with ID centrally positioned, ASD–ID, ID–CP, and CP more peripheral, and ID–EP and EP–ID closely related (Fig.2b). Gene membership for each cluster and the number of probands with variant in those genes are provided in Supplementary Tables 2 and 3.

**Fig. 1.**
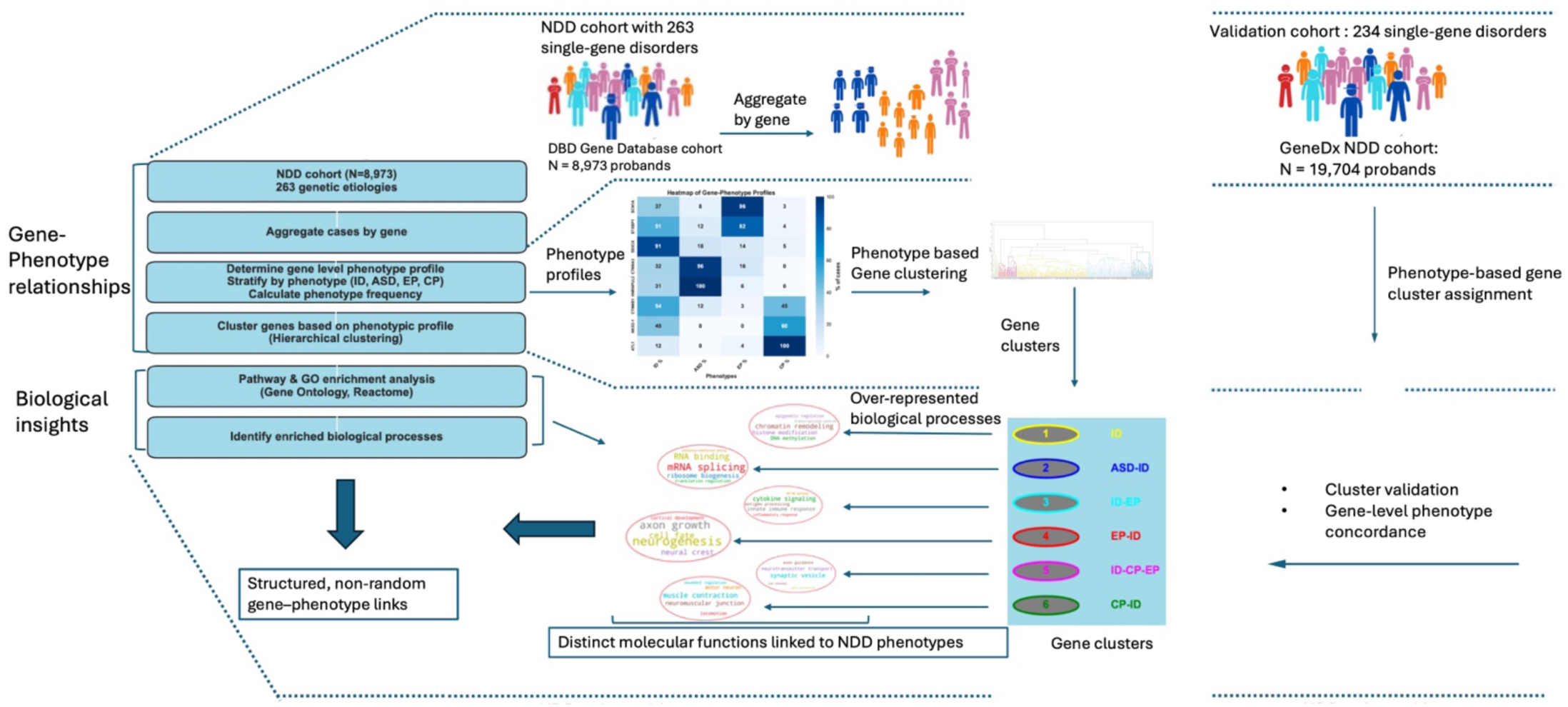
Study overview. We analyzed 8,973 probands with disease–causing variants in 263 high–confidence NDD genes and diagnoses intellectual disability and/or global developmental delay (ID), autism spectrum disorder (ASD), epilepsy (EP), or cerebral palsy (CP). For each gene, we generated a phenotype–frequency profiles and performed hierarchical, identifying six gene clusters. A validation cohort of 19,704 probands was used to validate cluster assignment of 234 overlapping genes, with cluster labels predicted by a Random Forest model trained on the discovery cohort. Validation assessed concordance of gene cluster assignments and cluster–level phenotype profiles across datasets. Cluster–specific Gene Ontology analyses were then performed to identify biological processes enriched within each cluster.

**Fig. 2.**
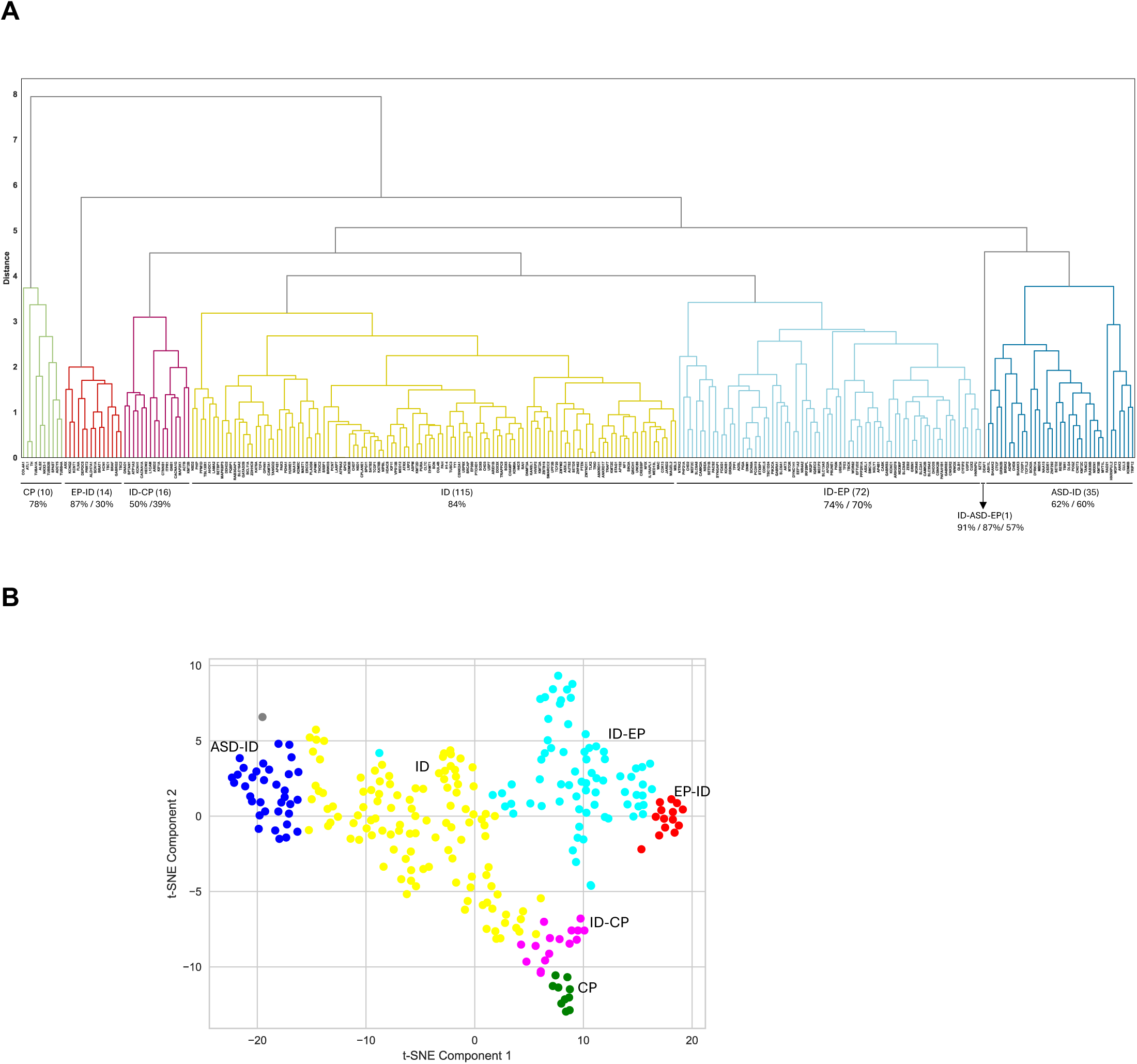
Phenotype–based Hierarchical clustering of NDD genes. **A** Hierarchical clustering of 263 NDD genes identified six distinct gene clusters. One gene (*DEAF1*) did not cluster and was excluded from downstream analyses. Each cluster is characterized by a unique distribution of phenotype frequencies. Cluster labels reflect the NDD phenotype(s) whose median frequency across genes within the cluster was at least 25%. Colors indicate cluster membership for visualization. **B** T–SNE projects gene–phenotype data into two dimensions, visualizing the six clusters identified by hierarchical clustering. Each point represents a gene, and colors indicate cluster membership. The number of genes for each cluster are CP (N=10); EP–ID (N=14); ID–CP (N=16); ID (N=115); ID–EP (N=72); ASD–ID (N=35). The single gene that did not cluster with any other genes is shown in grey

### Phenotypic profiles across gene clusters

To characterize phenotypic profiles across gene clusters, we summarized median disorder frequencies with 95% confidence intervals (Table 1). Phenotype profiles were generally consistent within clusters, with most genes showing similar profiles across ID, ASD, EP, and CP; variation was mainly attributable to low-frequency phenotypes (Fig.3a; Supplementary Fig. 3). As shown in Fig. 3A, clusters captured sets of genes with shared phenotypic profiles, underscoring the coherence of the cluster structure.

**Fig. 3.**
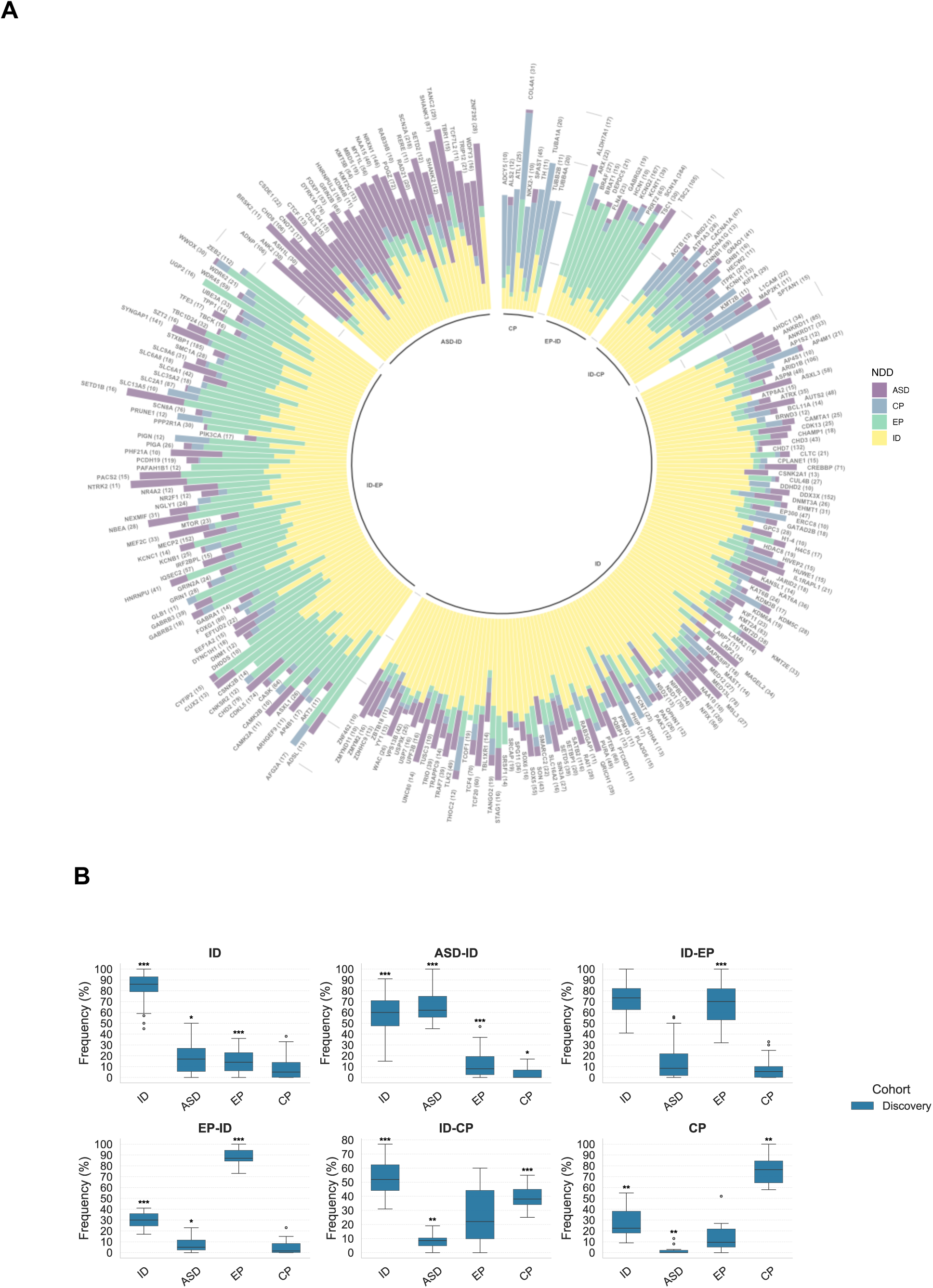
Phenotype profiles significantly differed across gene clusters. **A** Stacked bar plots show the relative frequency of each disorder across genes, grouped by gene cluster in the discovery dataset. **B** Boxplots show the distribution of disorder frequencies for genes within each cluster. Disorder frequencies within clusters were compared to the overall gene set using a two-sided one-sample Wilcoxon signed-rank test. The p–values were corrected for multiple hypothesis testing using the Benjamini–Hochberg procedure. *: p < 0.05; **: p < 0.01, ***: p < 0.001.

**Table 1.** Summary of gene–level phenotype frequencies across gene clusters in the discovery dataset^1^.

| NDDs | ID | ASD-ID | ID-EP | EP-ID | ID-CP | CP |
| --- | --- | --- | --- | --- | --- | --- |
| ID | 86 (85–89) | 60 (53–70) | 74 (70–78) | 30 (24–36) | 50 (44–62) | 22 (15–46) |
| ASD | 16 (11–20) | 62 (57–71) | 9 (5–13) | 5 (0–12) | 9 (5–12) | 0 (0–2) |
| EP | 14 (10–16) | 8 (6–14) | 70 (64–73) | 87 (84–94) | 22 (10–44) | 10 (4–25) |
| CP | 5 (4–7) | 0 (0–4) | 5 (2–7) | 1 (0–9) | 39 (35–45) | 78 (64–92) |

<sup>1</sup>For each cluster, values represent the median NDD phenotype frequency (%) across genes, with 95% confidence intervals shown in parentheses.

We next assessed whether each cluster exhibited disorder frequencies that differed from the cohort-wide profile. In the discovery cohort, cohort-wide frequencies were 75% for ID (IQR 57-88%), 14% for ASD (IQR 4-31%), 24% for EP (IQR 8-57%), and 6% for CP (IQR 0-15%). Using a two-sided one-sample Wilcoxon signed-rank test with Benjamini-Hochberg correction, we found clear and statistically robust cluster-specific signatures across all four disorders. These distinct phenotypic profiles were evident both in comparisons with the cohort-wide distribution (Fig. 3B) and in pairwise contrasts between clusters (Supplementary Fig.4).

**Fig. 4.**
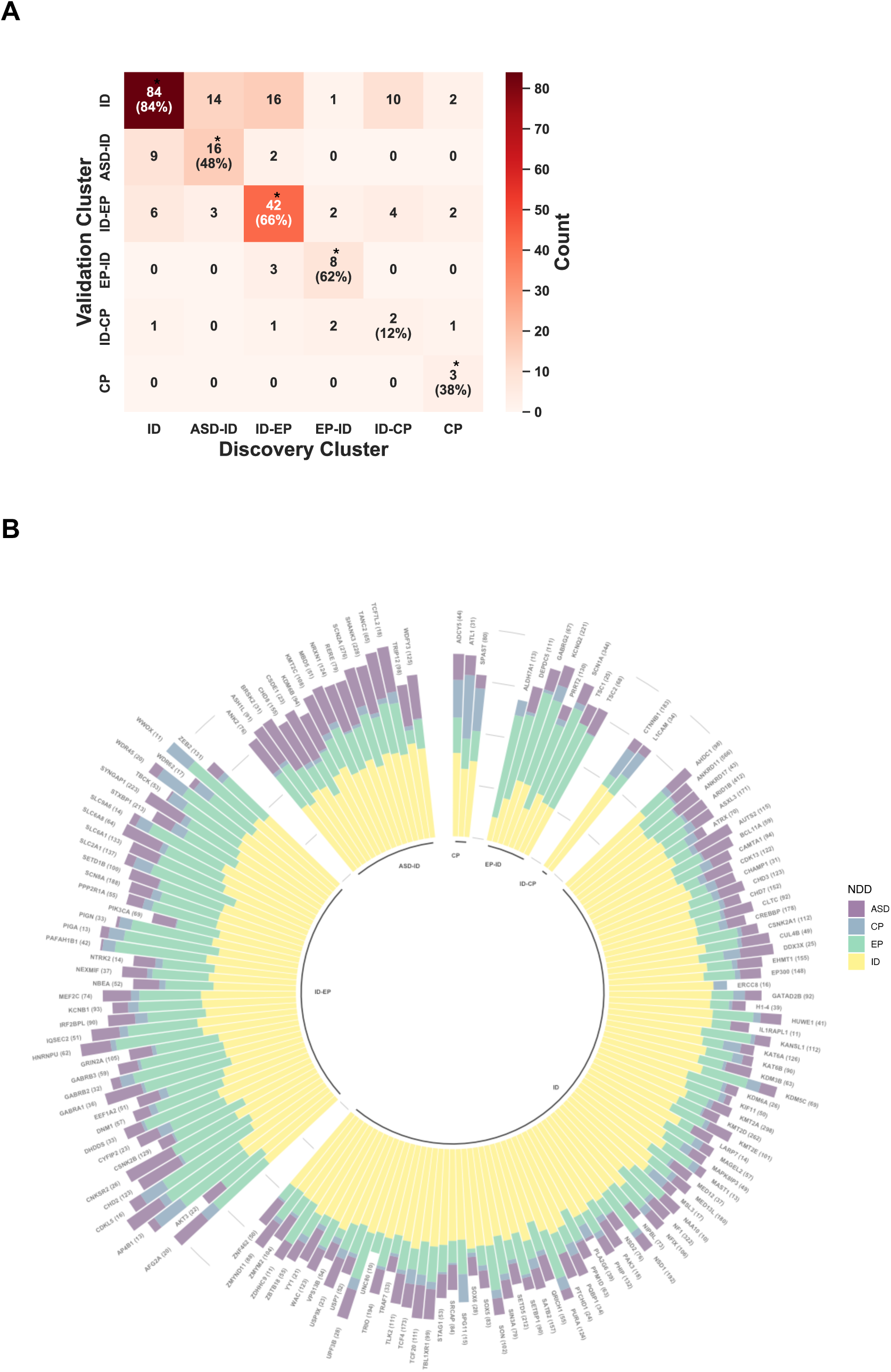
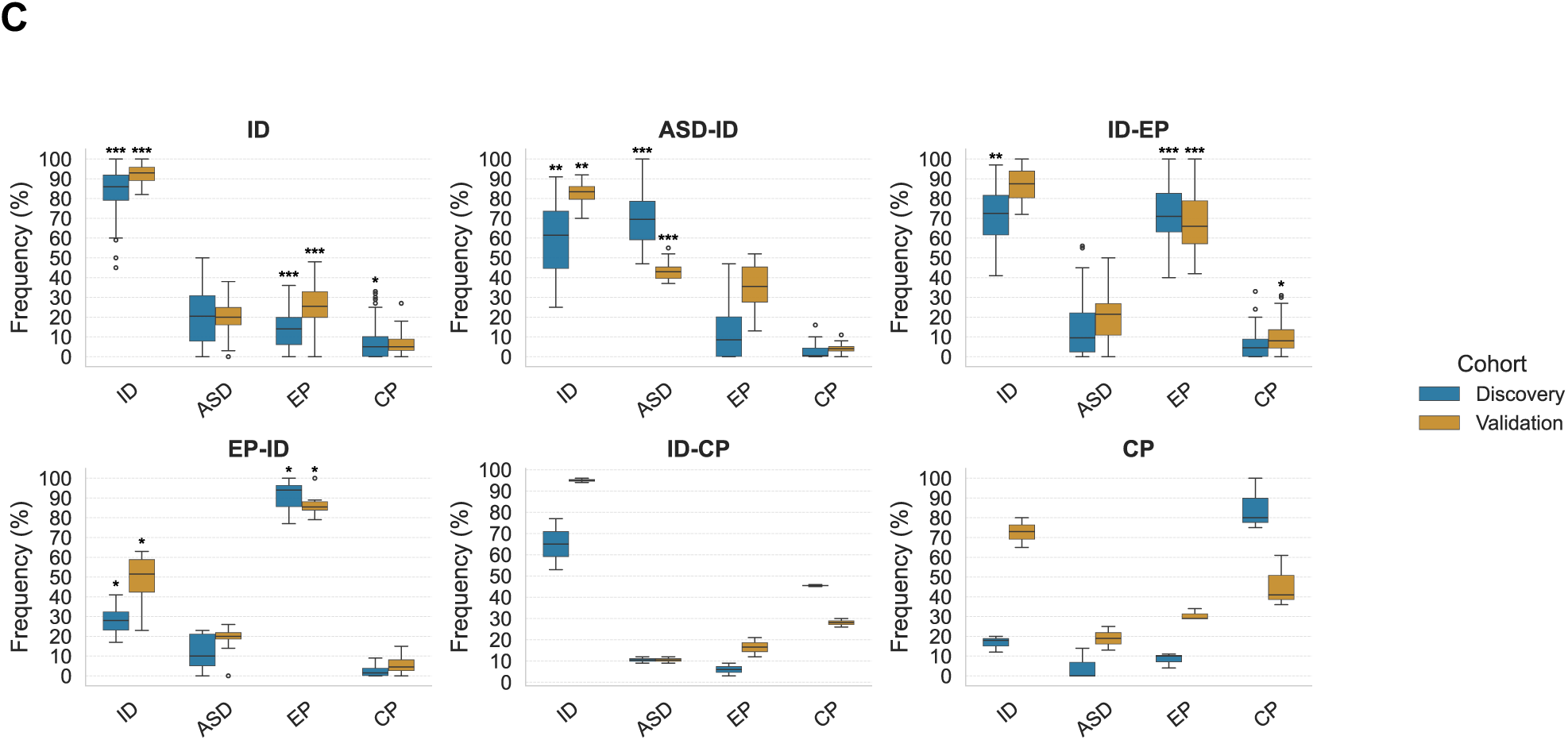
Supervised classification of clusters and identification of genes with concordant assignments between the discovery and validation datasets. **A** Confusion matrix showing Random Forest prediction of gene cluster assignments in the validation dataset. The matrix compares the model’s predicted validation labels (y–axis) against the discovery dataset labels (x–axis) for 234 genes. Color intensity corresponds to the number of genes per cell. Diagonal elements indicate genes assigned to the same cluster in both datasets (155/234 genes; 66%), with per-cluster percentage shown in parentheses. Clusters with more concordant assignments than expected by chance are indicated with asterisks (p <0.001; permutation test). Off–diagonal elements represent the number of genes assigned to different clusters in the two datasets. **B** Stacked bar plots show the relative frequency of NDD phenotypes across the 155 concordant genes in the validation dataset. **C** Gene-level disorder frequencies across clusters for the 155 concordant genes in both cohorts. Boxplots show distributions by cohort. For each disorder and cluster, deviation from the cohort-wide median were assessed using two-sided one-sample Wilcoxon signed-rank tests. P-values were corrected for multiple testing across all cluster–disorder comparisons using the Benjamini–Hochberg procedure. *: p < 0.05; **: p < 0.01, ***: p < 0.001.

The ID cluster, the largest cluster (N = 115), showed the strongest convergence around ID, with pronounced enrichment for ID (median 86%, p < 0.001), accompanied by significant depletion of EP (p < 0.001) and a modest increase in ASD frequency (16%, p < 0.05), while CP frequency did not differ from the cohort-wide distribution. The ASD–ID cluster (N = 35) exhibited the highest ASD frequency among all clusters (62%, p < 0.001), alongside a reduced, though still elevated ID frequency (60%, p < 0.01); EP and CP frequencies were significantly depleted (p < 0.001 and p < 0.01, respectively) relative to the cohort-wide median frequencies, supporting a shared ASD–ID phenotype distinct from EP or CP (Fig. 3b).

The ID–EP cluster (N = 72) showed high frequencies of both EP (70%, p < 0.001) and ID (74%), capturing genes with dual contributions to epileptic and cognitive phenotypes. While ID frequency remained high, it did not differ from the cohort median; ASD and CP frequencies were also unchanged. However, a small subset of genes within the cluster showed elevated ASD frequency, indicating overlap across ID, EP, and ASD (Supplementary Fig. 3c). In contrast, the EP–ID cluster (N = 14) showed the highest EP frequency (87%, p < 0.001), along with significantly reduced ID (30%, p < 0.001) and ASD frequencies (p < 0.05). This pattern is consistent with primarily epilepsy-driven genetic effects with only modest contribution to ID; CP frequency was low but not significantly different from the cohort (Fig. 3b).

The two CP-associated clusters revealed complementary phenotypic profile. The ID–CP cluster (N = 16) showed significantly elevated CP (39%, p < 0.001) alongside ID (50%, p < 0.001), but reduced ASD frequency (p < 0.01), while EP did not differ from the cohort median. Finally, the CP cluster (N = 10) showed the strongest CP enrichment (78%, p < 0.001) together with significant depletion of ID (22%, p < 0.001) and ASD (p < 0.0001). EP frequency was unchanged, highlighting a genetically distinct CP-associated cluster (Fig. 3b). Within these two CP-associated clusters, a small subset of genes showed elevated EP frequency, identifying genes that jointly influence ID, CP, and EP (Supplementary Fig. 3e-f).

Notably, ID was prevalent across all clusters (>20%), with frequency below 50% limited to the two smallest clusters, EP–ID and CP. Reciprocal patterns between ASD an CP were evident, with clusters enriched for CP consistently showed reduced ASD, whereas the cluster with high ASD frequency exhibited markedly low CP prevalence, suggesting distinct underlying genetic pathways (Table 1).

### Validation of gene clusters in a large independent NDD cohort

To evaluate the robustness of the gene clusters, we analyzed an independent validation cohort of 19,704 individuals from a commercial genetic testing laboratory, with causative variants in 234 of the 263 genes present in 94% (8,461/8,973) of discovery probands. We assessed two key aspects of validation: 1) stability of cluster membership across cohorts, meaning whether individual genes map to the same cluster in both datasets, and 2) the consistency of internal properties, including phenotype profiles and relative cluster sizes.

Across the 234 shared genes, median gene-level disorder frequencies differed between cohorts. ID and EP frequencies were higher in the validation cohort (ID: 90%, IQR 85–95%; EP: 36%, IQR 24–54%) than in the discovery cohort (ID: 75%, IQR 56–87%; EP: 24%, IQR 9–56%; Mann-Whitney U test, P < 0.0001). ASD frequencies also differed (validation: 21%, IQR 14–28%; discovery: 17%, IQR 5–33%; P < 0.05). Gene-level ASD frequencies in the discovery cohort were right-skewed, reflecting a subset of genes with elevated frequencies, whereas CP was low in both cohorts (Supplementary Fig. 5a). These cohort differences likely reflect ascertainment biases. The discovery cohort, aggregated from 1,150 published studies across diverse settings that included large cohorts, case series, and case reports, whereas the validation cohort comprised patients referred for clinical exome or genome sequencing based on NDD diagnosis, particularly ID. These distinct ascertainment strategies contributed to differences in disorder frequencies. Despite these differences in cohort composition, the overall gene clustering patterns were reproducible, supporting generalizability of cluster structure.

Using a result-based cluster validation strategy (Methods), we assigned the 234 shared genes in the validation dataset to six gene clusters. Cluster membership was predicted based on gene-level phenotype frequencies in the validation cohort using a Random Forest classifier. Of the 234 genes, 155 (66%) were assigned to the same cluster in both datasets –– significantly exceeding the 75.2 matches expected by chance (permutation test, p<0.0001). Of the 79 discordant genes, 43 were reassigned to the ID cluster in the validation dataset, driven by substantially higher ID frequencies relative to other disorders in that cohort (Fig. 4a). Notably, 26 of 79 discordant genes (those assigned to different cluster between the two datasets) shared the same two most frequent disorders in both datasets (Supplementary Table 5), indicating that differences in absolute rather than relative frequencies largely accounted for their discordant cluster assignments.

Evaluation of cluster membership concordance between the two cohorts using the Adjusted Rand Index yielded 0.322, indicating moderate but meaningful consistency in cluster structure across cohorts. Cluster–wise permutation tests further showed that five of the six clusters (ID, ASD–ID, ID–EP, EP–ID, and CP) exhibited significantly greater concordance than expected by chance (adjusted p<0.0001; Fig. 4a, Supplementary Table 6). The ID gene cluster, followed by the ID–EP, EP–ID, ASD–ID, and CP clusters, showed the largest number of concordant gene assignments, as shown in the diagonal elements of the confusion matrix (Fig.4a). In contrast, The ID–CP cluster showed considerable inconsistency, with most of its genes assigned to the ID cluster in the validation cohort (Fig. 4a).

Relative cluster sizes were also consistent across datasets. The ID cluster remained the largest (50%), followed by the ID–EP (∼25%), ASD–ID (∼15%), and EP–ID (∼10%) clusters, while the ID–CP and CP–ID clusters contained relatively few genes in both datasets (Supplementary Fig. 5b; Supplementary Tables 5 and 6). This close agreement in relative cluster size and composition across datasets provides further evidence that the underlying structures of the clusters is preserved, despite differences in absolute disorder frequencies.

Phenotype frequencies were highly similar between cohorts for the 234 shared genes and the 155 with concordant cluster assignments, particularly in the ID, ASD–ID, ID–EP, and EP–ID clusters (Fig. 4b-c). Among the 155 concordant genes, normalized disorder frequencies showed strong cross-cohort correlations, confirming stable gene-phenotype relationships and further supporting the reproducibility of internal cluster properties (Supplementary Fig. 6). Together, these results demonstrate that five of the six gene clusters (ID, ASD–ID, ID–EP, EP–ID, and CP) are reproducible across large, independently ascertained cohorts, supporting their robustness and generalizability. The ID–CP cluster did not replicate, likely reflecting its small size and sensitivity to cohort ascertainment.

Validation of gene clusters using 149 genes in a subset of the validation cohort that excluded any potential overlap with the discovery cohort (Methods; Supplementary Table 7) yielded phenotype profiles consistent with those observed in the full cohort, with concordant cluster assignment for 99 of 149 genes (60%) (permutation test, p < 0.0001; Supplementary Table 8; Extended Data Fig.7).

### Biological processes and pathways underpinning NDD phenotypes

To investigate the biological functions represented within the six gene clusters from the discovery dataset, we performed Gene Ontology (GO) enrichment analysis on the 262 clustered genes. Across clusters, 264 unique GO biological processes were significantly enriched and supported by at least two genes per cluster (Supplementary Table 9). Distinct enrichment patterns and magnitudes across clusters captures a different set of biological themes, reflecting the unique phenotypic profiles identified earlier.

Enriched GO processes (Supplementary Table 9), including the top 10 shown in Fig. 5a, highlight strong cluster-specific biological signatures, and network visualizations illustrate the genes driving these enrichments (Supplementary Fig. 8). Pairwise comparisons revealed minimal GO term overlap and low semantic similarity between clusters (0.3-0.5), consistent with limited functional relatedness and distinct biological roles. The highest semantic similarity occurred between ASD–ID and ID–EP, and between ID–EP and EP–ID clusters (similarity=0.51), while the lowest was between the ID and EP–ID clusters (similarity=0.35; Supplementary Table 10).

**Fig. 5.**
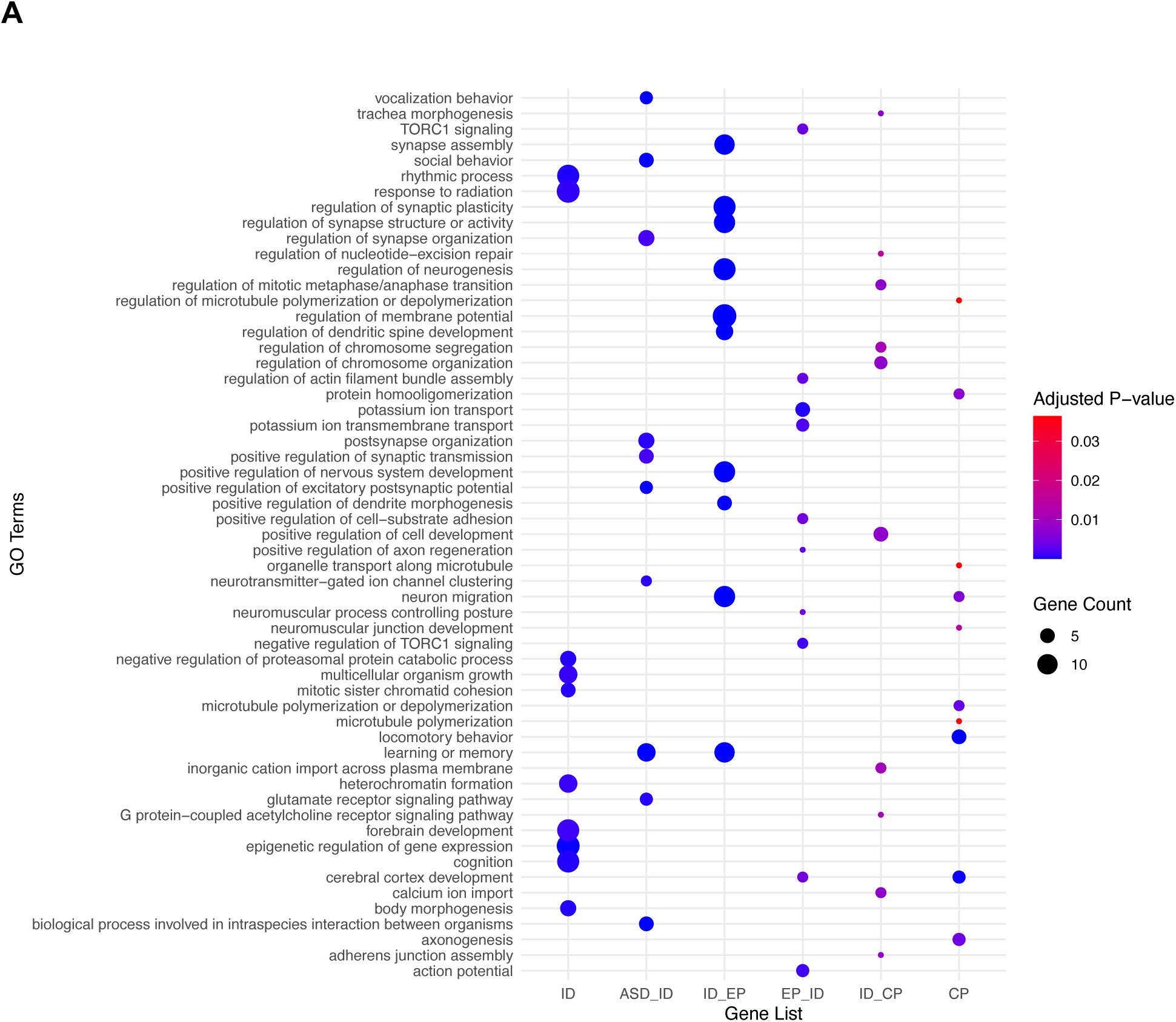
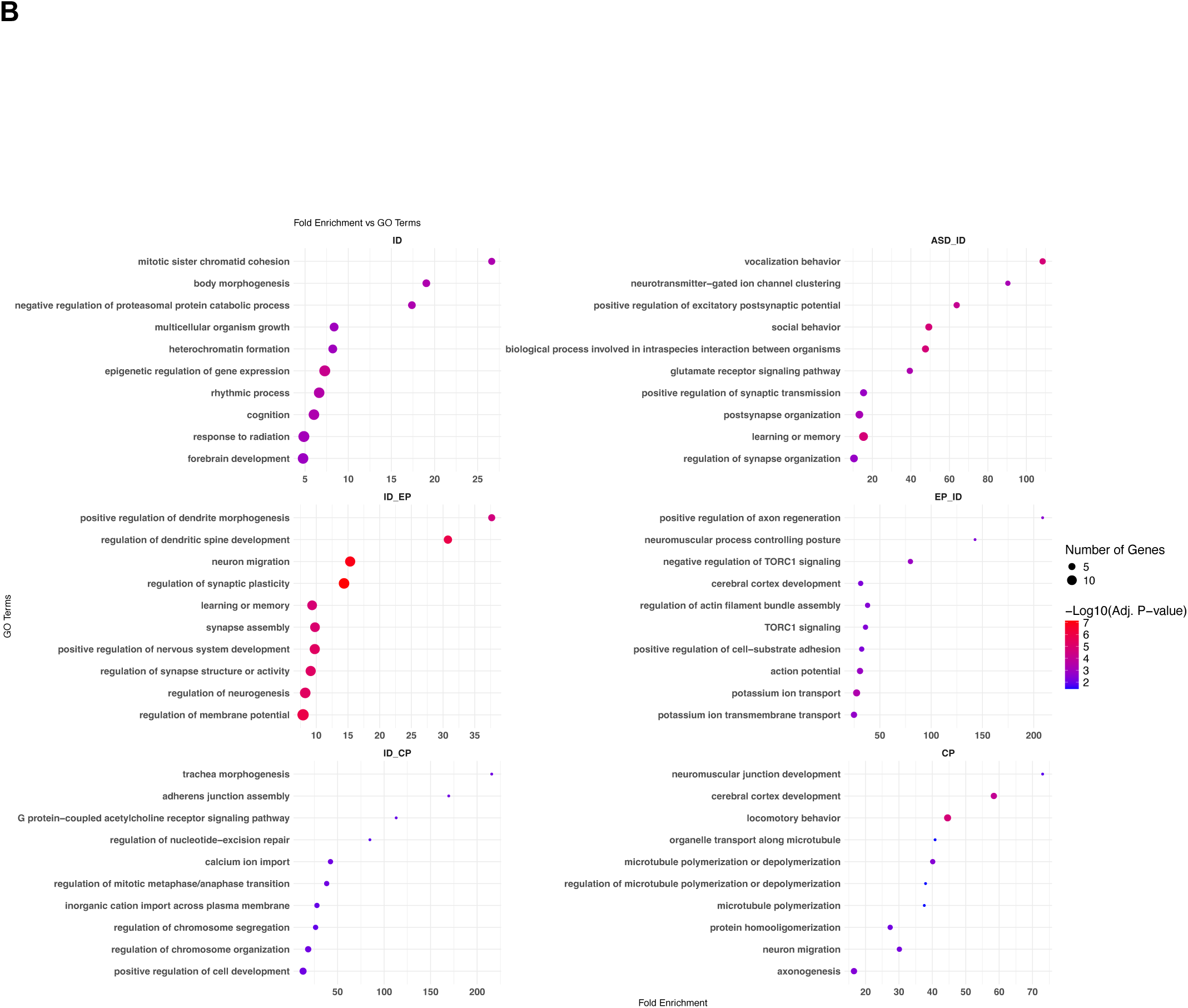
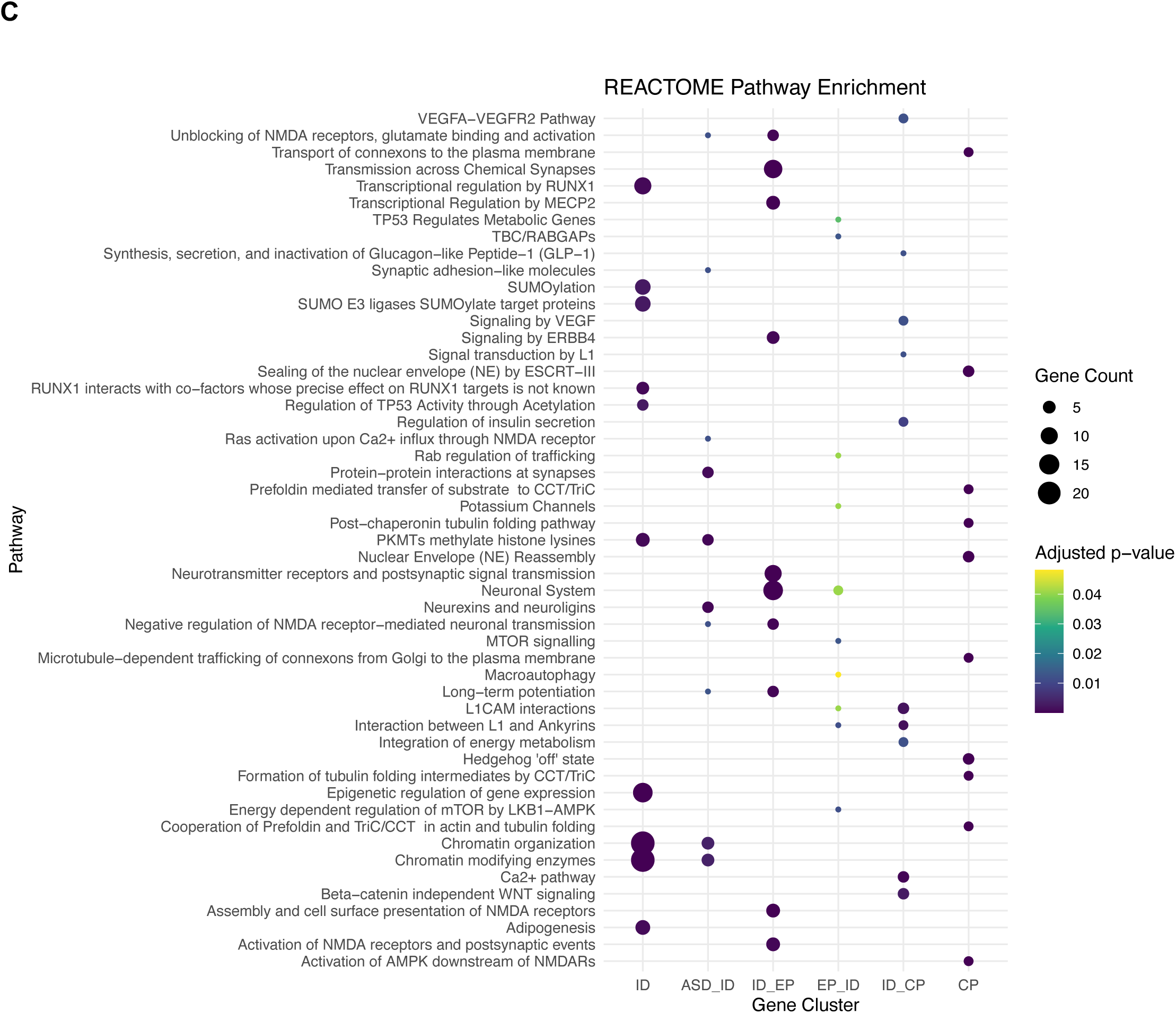
Distinct GO biological processes and pathways are enriched within each gene cluster. **A** The top 10 enriched biological processes with adjusted p–value < 0.05 are plotted. **B** Fold enrichment of enriched biological process in each cluster. **C** The top 10 significantly enriched Reactome pathways colored by adjusted p–values (<0.05). Gene clusters are shown on the x–axis. The size of each dot corresponds to the number genes in a pathway.

Although most enriched processes were cluster-specific, 12% (31 of 264) were shared, indicating common biological processes contributing to multiple NDD phenotypes (Supplementary Table 9). Shared processes often showed markedly different enrichment strength across clusters. For example, glutamate receptor signaling pathway (GO:0007215) was enriched 39-fold in ASD–ID and 19-fold in ID–EP, while locomotor behavior (GO:0007626) showed 45-, 12-, and 6-fold enrichment (FE) in CP, EP–ID, and ID–EP clusters, respectively (Supplementary Table 9). Notably, ∼80% of genes in each cluster contributed to enriched processes (Supplementary Table 11), indicating strong functional coherence within clusters.

Each cluster showed distinct biological processes enrichment (Fig.5a, Supplementary Table 9). The ID cluster was enriched for chromatin organization, nervous system development, and gene regulation, with top terms including mitotic sister chromatid cohesion (FE=26.6, p=4.5×10^-4^) and body morphogenesis (FE=19.0, p=4.2×10^-4^). The ASD–ID cluster highlighted synaptic organization and behavioral regulation, notably vocalization behavior (FE=108.5, p=1.8×10^-5^) and neurotransmitter-gated ion channel clustering (FE=90.4, p=6.0×10^-4^). The ID–EP cluster was enriched for synaptic activity and neuronal development, including dendrite morphogenesis (FE=37.7, p=2.6×10^-5^) and neuron migration (FE=15.3, p=1.2×10^-7^). EP–ID showed strong enrichment in positive regulation of axon regeneration (FE=208.6, p=2.9×10^-3^), neuromuscular processes controlling posture (FE=142.8, p=4.3×10^-3^), and negative regulation of TORC1 signaling (FE=79.8, p=1.4×10^-3^). The ID–CP cluster was associated with cell development, morphogenesis, and DNA repair, with top terms such as trachea morphogenesis (FE=215.8, p=7.6×10^-3^). Finally, the CP cluster was enriched for neuromuscular junction development (FE=73.0, p=0.01), cerebral cortex development (FE=58.4, p=9.5×10^-5^), and locomotory behavior (FE=44.6, p=1.8×10^-5^). Together, these enrichments reveal that each cluster is linked to a distinct biological architecture consistent with its phenotypic signatures (Fig. 5b, Supplementary Table 9).

REACTOME pathway analysis identified 234 significantly enriched pathways across the six clusters, each supported by at least two genes, with 42 pathways (18%) shared between clusters (Supplementary Table 12). The top ten pathways highlight distinct functional modules represented by genes within each cluster, (Fig.5c, Supplementary Fig. 9). Overall pathway overlap was limited, consistent with divergent biological mechanisms across clusters. However, ASD–ID and ID–EP showed substantial overlap, particularly in synaptic and neuronal pathways, including neurexins, neuroligins, and NMDA receptor signaling (Supplementary Table 12). Of the 15 pathways enriched in ASD–ID, 14 were also enriched in other clusters, most predominantly in ID–EP, although they ranked lower outside ASD–ID’s top pathways (Supplementary Table 12). The only ASD–ID–specific pathway was the deactivation of the beta-catenin transactivating complex, linked to *CHD8* and *TCF7L2,* highlighting a regulatory process unique to ASD–ID genes. Together, these results suggest that both shared and cluster-specific pathways contribute to differentiation of NDD phenotypes, reflecting convergent and divergent biological mechanisms across gene sets.

We next assessed whether these biological themes persisted when limiting analyses on the 155 genes with concordant cluster assignments between the discovery and validation datasets (Supplementary Fig. 10). Although this subset yielded fewer enriched GO terms (143 vs. 264), the core biological themes and low semantic similarity of GO terms between clusters were preserved (Supplementary Tables 13 and14). Reactome pathway enrichment also largely recapitulated the full-set results, though no pathways reached significance in the CP–ID or CP clusters, which contained too few genes (Supplementary Table 15).

## Discussion

By clustering a large set of high-confidence NDD genes within a cross-disorder cohort of 263 monogenic NDDs, we identified six gene clusters with distinct phenotypic profiles, providing a structured view of how pathogenic variants drive both shared and divergent neurodevelopmental outcomes. These clusters include genes that primarily influence cognition, jointly affect ASD and ID, or epilepsy and ID, or motor/CP phenotypes, extending findings from single diagnosis-ascertained cohorts. Unlike single-disorder designs that infer gene effects from ASD-, EP- or developmental delay-ascertained cohorts without considering comorbid diagnoses [5, 11, 23, 26], our cross-disorder approach quantifies gene-level frequencies across all four conditions, enabling accurate detection of genes that influence multiple disorders.

The phenotypic distinctions across clusters aligned with established gene-phenotype relationships. Our analyses revealed a large, ID gene cluster characterized by intellectual disability/developmental delay, with autism and epilepsy occurring infrequently. This phenotypic profile likely reflects the stabilizing effect of large-scale aggregation of gene-level phenotypes and aligns with prior reports that these genes are enriched in developmental delay rather than ASD-ascertained cohort [6]. Consistent with frequent co-occurrence of ASD and ID [17, 21, 51, 52], we observed a robust ASD-ID gene cluster but no ASD-only cluster. Other clusters mapped to well-established gene-phenotype relationships: the ID–EP cluster includes developmental and epileptic encephalopathy genes, (e.g., *CDKL5*, *FOXG1*, *STXBP1*, *SCN8A*) [7], whereas the EP–ID cluster captured epilepsy-exclusive phenotypes (*PRRT2*, *GABRG2*) [7, 8]. The ID–CP and CP clusters were enriched for high-confidence CP-associated genes, including *CTNNB1, SPAST, COL4A1* and tubulin genes (e.g., *TUBA1A*, *TUBB2B*, and *TUBB4A)* [10, 18, 53]. Together, these patterns reinforce the robustness of our clustering framework for distinguishing shared and distinct signatures across NDD genes.

The replication of five of six gene clusters (ID, ASD–ID, ID–EP, EP–ID, and CP) in an independent cohort supports their robustness, and indicates that many genes converge on recurring phenotypic profiles, consistent with the view that monogenic causes transcend categorical diagnoses and that shared genetic etiologies contribute to overlapping developmental phenotypes [54–56]. Using cross-disorder probands allowed us to refine phenotypic profiles across diagnostic boundaries, which was replicated across cohorts despite differences in ascertainment. Although most clusters replicated well, additional work is needed to clarify whether the ID–CP and CP–ID clusters represent distinct groups or a single CP-enriched cluster.

Ontology-based enrichment revealed distinct functional themes across clusters. The ID cluster showed strong enrichment for chromatin regulation pathways, including genes involved in histone methylation (*KMT2D*, *KMT2A*, or *NSD1*), histone acetylation (*CREBBP*, *EP300*, *KAT6A*), *SWI/SNF* chromatin-remodeling (*ARID1B*, *SMARCC2*, or *CHD3*), and chromatin architecture (*ATRX*, *NIPBL*, or *SATB2*) [57–59]. In contrast, the ASD–ID cluster was enriched for synapse development and maintenance [16, 22, 35] and, the EP–ID cluster for synaptic transmission and ion transport aligning with past reports [26]; our analysis also found new enrichments, such as actin-mediated cell contraction. The CP and ID–CP clusters showed expected enrichment for locomotor-related processes, mirroring findings from other studies [60, 61]. Notably, locomotory behavior enrichment across EP–ID and ID–EP clusters highlight epilepsy-associated genes (*MECP2, MTOR, NR4A2*, *PAFAH1B1*, *SCN2A*, *TSC2*) that also contribute to motor dysfunction, pointing to shared neurobiological mechanisms underlying seizure and motor phenotypes [62–66].

Several limitations warrant consideration. First, NDD diagnoses in the discovery cohort are curated from published literature and may underestimate true phenotype frequencies due to incomplete reporting, and CP cases were underrepresented because genomic studies of CP are relatively recent. ID may be overrepresented given its longstanding emphasis in genetic studies. Although minor sample overlap between cohorts cannot be fully excluded, sensitivity analyses restricted to known non-overlapping validation cases produced similar results. We also excluded emerging genes to focus on well-established NDD genes and did not distinguish between missense and loss-of-function variants, despite some genes showing variant-specific phenotypic differences. Additionally, we did not assess phenotype frequencies by age, sex, or inheritance, which may influenced phenotypic expression pattern. Nonetheless, the mechanistic and phenotypic convergence across numerous NDD genes suggests that the enriched processes identified here represent core biological functions. Future work incorporating additional datasets, broader NDD phenotypes and external phenotypic validation will help refine and test the robustness of these clusters.

By identifying groups of genes that converge on similar phenotypic profiles, our framework can improve diagnostic interpretation, support gene-informed prognostic counseling, and help prioritize mechanistically coherent gene sets for future therapeutic research, addressing a major barrier to randomized clinical trials posed by the rarity of individual genetic etiologies. Gene-level phenotypic and functional annotations may further aid in variant prioritization, particularly for variants of uncertain significance, and pathway based screening can help reveal novel NDD genes in unsolved cases [54]. Ultimately, recognizing clusters with distinct phenotypic signatures will clarify mechanisms by which pathogenic genomic variants lead to similar or divergent clinical NDD trajectories.

## Data Availability

All data produced in the present work are contained in the manuscript

## Acknowledgements

This research was supported by the National Institutes of Mental Health under award number R01MH074090 to David H. Ledbetter and Christa L. Martin and the Eunice Kennedy Shriver National Institute of Child Health and Human Development under award number R01HD104938 to Scott M. Myers. Ingo Helbig is supported by the National Institute for Neurological Disorders and Stroke (R01 NS131512, R01 NS127830) and the Hartwell Foundation (Individual Biomedical research Award).

Ingo Helbig received support through the German Research Foundation (DFG/FNR INTER Research Unit FOR2715 (He5415/7-1 and He5415/7-2).

## Conflict of Interests

David Ledbetter is a scientific consultant to Nest Genomics, Inc. and MyOme, Inc.

Bobbi McGivern and Zhancheng Zhang are employees of and may own stock in GeneDx.

## Notes

### Author Declarations

This study includes two cohorts. The first, the discovery cohort, was identified from a publicly available database that aggregates genotype and phenotype information from published articles. The database is accessible at: https://dbd.geisingeradmi.org/. The second cohort, the validation cohort, includes individuals who underwent clinical exome or genome sequencing at GeneDx. This study was conducted under a GeneDx research protocol approved by the Western Institutional Review Board (Study Number 1169768; WIRB Pro Number 20162523), which permitted a waiver of consent.

### Summary of Updates

The fonts used in the original text have been updated to correct character display issues, and the figures have been revised to enhance their resolution.

## References

1. Lazar SM, Svoboda M, Risen S, Myers SM. Patterns of Typical and Atypical Neurodevelopment. In: Ismail FY, Accardo PJ, Shapiro BK, editors. Capute & Accardo’s Neurodevelopmental Disabilities in Infancy and Childhood, vol. 2025, London: Elsevier. p. 13–37.

2. Moreno-De-Luca A, Myers SM, Challman TD, Moreno-De-Luca D, Evans DW, Ledbetter DH. Developmental brain dysfunction: revival and expansion of old concepts based on new genetic evidence. Lancet Neurol. 2013;12:406–414.

3. Kaplanis J, Samocha KE, Wiel L, Zhang Z, Arvai KJ, Eberhardt RY, et al. Evidence for 28 genetic disorders discovered by combining healthcare and research data. Nature. 2020. October 14, 2020. 10.1038/s41586-020-2832-5.

4. Hamanaka K, Miyake N, Mizuguchi T, Miyatake S, Uchiyama Y, Tsuchida N, et al. Large-scale discovery of novel neurodevelopmental disorder-related genes through a unified analysis of single-nucleotide and copy number variants. Genome Med. 2022;14:40.

5. Wang T, Kim CN, Bakken TE, Gillentine MA, Henning B, Mao Y, et al. Integrated gene analyses of de novo variants from 46,612 trios with autism and developmental disorders. Proc Natl Acad Sci U S A. 2022;119:e2203491119.

6. Fu JM, Satterstrom FK, Peng M, Brand H, Collins RL, Dong S, et al. Rare coding variation provides insight into the genetic architecture and phenotypic context of autism. Nat Genet. 2022. August 18, 2022. 10.1038/s41588-022-01104-0.

7. Heyne HO, Singh T, Stamberger H, Jamra A, Caglayan R, Craiu H, et al. De novo variants in neurodevelopmental disorders with epilepsy. Nat Genet. 2018;50:1048–1053.

8. Oliver KL, Scheffer IE, Bennett MF, Grinton BE, Bahlo M, Berkovic SF. Genes4Epilepsy: An epilepsy gene resource. Epilepsia. 2023;64:1368–1375.

9. Wang Y, Xu Y, Zhou C, Cheng Y, Qiao N, Shang Q, et al. Exome sequencing reveals genetic heterogeneity and clinically actionable findings in children with cerebral palsy. Nat Med. 2024;30:1395–1405.

10. van Eyk CL, Fahey MC, Gecz J. Redefining cerebral palsies as a diverse group of neurodevelopmental disorders with genetic aetiology. Nat Rev Neurol. 2023;19:542–555.

11. Zhang Y, Wang R, Liu Z, Jiang S, Du L, Qiu K, et al. Distinct genetic patterns of shared and unique genes across four neurodevelopmental disorders. Am J Med Genet B Neuropsychiatr Genet. 2021;186:3–15.

12. Leblond CS, Le T-L, Malesys S, Cliquet F, Tabet A-C, Delorme R, et al. Operative list of genes associated with autism and neurodevelopmental disorders based on database review. Mol Cell Neurosci. 2021;113:103623.

13. Bonti E, Zerva IK, Koundourou C, Sofologi M. The high rates of comorbidity among neurodevelopmental disorders: Reconsidering the clinical utility of distinct diagnostic categories. J Pers Med. 2024;14:300.

14. Mitchell KJ. The genetics of neurodevelopmental disease. Curr Opin Neurobiol. 2011;21:197–203.

15. Coe BP, Stessman HAF, Sulovari A, Geisheker MR, Bakken TE, Lake AM, et al. Neurodevelopmental disease genes implicated by de novo mutation and copy number variation morbidity. Nat Genet. 2019;51:106–116.

16. Satterstrom FK, Kosmicki JA, Wang J, Breen MS, De Rubeis S, An J-Y, et al. Large-Scale Exome Sequencing Study Implicates Both Developmental and Functional Changes in the Neurobiology of Autism. Cell. 2020;180:568–584.e23.

17. Myers SM, Challman TD, Bernier RA, Bourgeron T, Chung WK, Constantino JN, et al. Insufficient evidence for “autism-specific” genes. Am J Hum Genet. 2020;106:587–595.

18. Moreno-De-Luca A, Millan F, Pesacreta DR, Elloumi HZ, Oetjens MT, Teigen C, et al. Molecular Diagnostic Yield of Exome Sequencing in Patients With Cerebral Palsy. JAMA. 2021;325:467–475.

19. Willsey AJ, Morris MT, Wang S, Willsey HR, Sun N, Teerikorpi N, et al. The Psychiatric Cell Map Initiative: A Convergent Systems Biological Approach to Illuminating Key Molecular Pathways in Neuropsychiatric Disorders. Cell. 2018;174:505–520.

20. Manoli DS, State MW. Autism spectrum disorder genetics and the search for pathological mechanisms. Am J Psychiatry. 2021;178:30–38.

21. Willsey HR, Willsey AJ, Wang B, State MW. Genomics, convergent neuroscience and progress in understanding autism spectrum disorder. Nat Rev Neurosci. 2022;23:323–341.

22. Asif M, Martiniano HFMC, Marques AR, Santos JX, Vilela J, Rasga C, et al. Identification of biological mechanisms underlying a multidimensional ASD phenotype using machine learning. Transl Psychiatry. 2020;10:43.

23. Ganesan S, Ruggiero SM, Parthasarathy S, Galer PD, Lewis-Smith D, McSalley I, et al. Phenotypic analysis of 11,125 trio exomes in neurodevelopmental disorders. 2025.

24. Lewis SA, Shetty S, Wilson BA, Huang AJ, Jin SC, Smithers-Sheedy H, et al. Insights from genetic studies of cerebral palsy. Front Neurol. 2020;11:625428.

25. Magielski J, Mcsalley I, Parthasarathy S, Mckee J, Ganesan S, Helbig I. Advances in big data and omics: Paving the way for discovery in childhood epilepsies. Curr Probl Pediatr Adolesc Health Care. 2024;54.

26. Chow J, Jensen M, Amini H, Hormozdiari F, Penn O, Shifman S, et al. Dissecting the genetic basis of comorbid epilepsy phenotypes in neurodevelopmental disorders. Genome Med. 2019;11:65.

27. Doshi-Velez F, Ge Y, Kohane I. Comorbidity clusters in autism spectrum disorders: an electronic health record time-series analysis. Pediatrics. 2014;133:e54–63.

28. Pérez-Cano L, Boccuto L, Sirci F, Hidalgo JM, Valentini S, Bosio M, et al. Characterization of a clinically and biologically defined subgroup of patients with autism spectrum disorder and identification of a tailored combination treatment. Biomedicines. 2024;12:991.

29. Stevens E, Dixon DR, Novack MN, Granpeesheh D, Smith T, Linstead E. Identification and analysis of behavioral phenotypes in autism spectrum disorder via unsupervised machine learning. Int J Med Inform. 2019;129:29–36.

30. Narita A, Nagai M, Mizuno S, Ogishima S, Tamiya G, Ueki M, et al. Clustering by phenotype and genome-wide association study in autism. Transl Psychiatry. 2020;10:290.

31. Yin L, Chau CKL, Sham P-C, So H-C. Integrating clinical data and imputed transcriptome from GWAS to uncover complex disease subtypes: Applications in psychiatry and cardiology. Am J Hum Genet. 2019;105:1193–1212.

32. Lagorce D, Lebreton E, Matalonga L, Hongnat O, Chahdil M, Piscia D, et al. Phenotypic similarity-based approach for variant prioritization for unsolved rare disease: a preliminary methodological report. Eur J Hum Genet. 2024;32:182–189.

33. Díaz-Santiago E, Jabato FM, Rojano E, Seoane P, Pazos F, Perkins JR, et al. Phenotype-genotype comorbidity analysis of patients with rare disorders provides insight into their pathological and molecular bases. PLoS Genet. 2020;16:e1009054.

34. McGuirl MR, Smith SP, Sandstede B, Ramachandran S. Detecting shared genetic architecture among multiple phenotypes by hierarchical clustering of gene-level association statistics. Genetics. 2020;215:511–529.

35. Emberti Gialloreti L, Enea R, Di Micco V, Di Giovanni D, Curatolo P. Clustering Analysis Supports the Detection of Biological Processes Related to Autism Spectrum Disorder. Genes. 2020;11.

36. Di Giovanni D, Enea R, Di Micco V, Benvenuto A, Curatolo P, Emberti Gialloreti L. Using machine learning to explore shared genetic pathways and possible endophenotypes in autism spectrum disorder. Genes (Basel). 2023;14:313.

37. Grotzinger AD, Werme J, Peyrot WJ, Frei O, de Leeuw C, Bicks LK, et al. Mapping the genetic landscape across 14 psychiatric disorders. Nature. 2026;649:406–415.

38. Ciulkinyte A, Mountford HS, Fontanillas P, 23andMe Research Team, Bates TC, Martin NG, et al. Genetic neurodevelopmental clustering and dyslexia. Mol Psychiatry. 2025;30:140–150.

39. Romanovsky E, Choudhary A, Peles D, Abu-Akel A, Stern S. Uncovering convergence and divergence between autism and schizophrenia using genomic tools and patients’ neurons. Mol Psychiatry. 2025;30:1019–1028.

40. Litman A, Sauerwald N, Green Snyder L, Foss-Feig J, Park CY, Hao Y, et al. Decomposition of phenotypic heterogeneity in autism reveals underlying genetic programs. Nat Genet. 2025:1–9.

41. Qiao J, Jiang L, Cai L, Chang M, Wang C, Zhao R, et al. Shared genetic architecture contributes to risk of major cardiovascular diseases. Nat Commun. 2025;16:8368.

42. Wang T, Hoekzema K, Vecchio D, Wu H, Sulovari A, Coe BP, et al. Large-scale targeted sequencing identifies risk genes for neurodevelopmental disorders. Nat Commun. 2020;11:4932.

43. Ullmann T, Hennig C, Boulesteix A-L. Validation of cluster analysis results on validation data: A systematic framework. Wiley Interdiscip Rev Data Min Knowl Discov. 2022;12:e1444.

44. Kapp AV, Tibshirani R. Are clusters found in one dataset present in another dataset? Biostatistics. 2007;8:9–31.

45. Yu G, Wang L-G, Han Y, He Q-Y. clusterProfiler: an R package for comparing biological themes among gene clusters. OMICS. 2012;16:284–287.

46. Wu T, Hu E, Xu S, Chen M, Guo P, Dai Z, et al. clusterProfiler 4.0: A universal enrichment tool for interpreting omics data. Innovation (Camb). 2021;2:100141.

47. Yu G, Wang L-G, Yan G-R, He Q-Y. DOSE: an R/Bioconductor package for disease ontology semantic and enrichment analysis. Bioinformatics. 2015;31:608–609.

48. Yu G. Gene ontology semantic similarity analysis using GOSemSim. Methods Mol Biol. 2020;2117:207–215.

49. Yu G, Li F, Qin Y, Bo X, Wu Y, Wang S. GOSemSim: an R package for measuring semantic similarity among GO terms and gene products. Bioinformatics. 2010;26:976–978.

50. Yu G, He Q-Y. ReactomePA: an R/Bioconductor package for reactome pathway analysis and visualization. Mol Biosyst. 2016;12:477–479.

51. Shaw KA, Williams S, Patrick ME, Valencia-Prado M, Durkin MS, Howerton EM, et al. Prevalence and early identification of autism spectrum disorder among children aged 4 and 8 years - autism and Developmental Disabilities Monitoring Network, 16 sites, United States, 2022. MMWR Surveill Summ. 2025;74:1–22.

52. Savatt JM, Myers SM. Genetic Testing in Neurodevelopmental Disorders. Front Pediatr. 2021;9:526779.

53. Gonzalez-Mantilla PJ, Hu Y, Myers SM, Finucane BM, Ledbetter DH, Martin CL, et al. Diagnostic yield of exome sequencing in cerebral palsy and implications for genetic testing guidelines: A systematic review and meta-analysis. JAMA Pediatr. 2023;177:472–478.

54. Deciphering Developmental Disorders Study. Large-scale discovery of novel genetic causes of developmental disorders. Nature. 2015;519:223–228.

55. Gonzalez-Mantilla AJ, Moreno-De-Luca A, Ledbetter DH, Martin CL. A Cross-Disorder Method to Identify Novel Candidate Genes for Developmental Brain Disorders. JAMA Psychiatry. 2016;73:275–283.

56. Andrews T, Meader S, Vulto-van Silfhout A, Taylor A, Steinberg J, Hehir-Kwa J, et al. Gene networks underlying convergent and pleiotropic phenotypes in a large and systematically-phenotyped cohort with heterogeneous developmental disorders. PLoS Genet. 2015;11:e1005012.

57. Iwase S, Bérubé NG, Zhou Z, Kasri NN, Battaglioli E, Scandaglia M, et al. Epigenetic etiology of intellectual disability. J Neurosci. 2017;37:10773–10782.

58. Chiurazzi P, Pirozzi F. Advances in understanding - genetic basis of intellectual disability. F1000Res. 2016;5:599.

59. Marti M, Millan MIP, Young JI, Walz K. Intellectual disability, the long way from genes to biological mechanisms. Journal of Translational Genetics and Genomics. 2020. 2020. 10.20517/jtgg.2020.10.

60. Zhu Q, Ni Y, Wang J, Yin H, Zhang Q, Zhang L, et al. Identification of pathways and genes associated with cerebral palsy. Genes Genomics. 2018;40:1339–1349.

61. Jin SC, Lewis SA, Bakhtiari S, Zeng X, Sierant MC, Shetty S, et al. Mutations disrupting neuritogenesis genes confer risk for cerebral palsy. Nat Genet. 2020;52:1046–1056.

62. Gonzales ML, LaSalle JM. The role of MeCP2 in brain development and neurodevelopmental disorders. Curr Psychiatry Rep. 2010;12:127–134.

63. Bockaert J, Marin P. MTOR in brain physiology and pathologies. Physiol Rev. 2015;95:1157–1187.

64. Gabaldon-Albero A, Mayo S, Martinez F. NR4A2 as a novel target gene for developmental and epileptic encephalopathy: A systematic review of related disorders and therapeutic strategies. Int J Mol Sci. 2024;25:5198.

65. de Wit M-CY, de Rijk-van Andel J, Halley DJ, Poddighe PJ, Arts WFM, de Coo IFM, et al. Long-term follow-up of type 1 lissencephaly: survival is related to neuroimaging abnormalities. Dev Med Child Neurol. 2011;53:417–421.

66. Turner TJ, Zourray C, Schorge S, Lignani G. Recent advances in gene therapy for neurodevelopmental disorders with epilepsy. J Neurochem. 2021;157:229–262.

