## Supplementary Figures for "Shared and distinct phenotypic profiles among neurodevelopmental disorder genes"

### Supplementary Information

#### Supplementary Materials and Methods

##### Identification of study cohorts and gene sets

###### *Discovery cohort*

The discovery cohort was identified from the Geisinger Developmental Brain Disorder (DBD) Gene Database (https://dbd.geisingeradmi.org/) [1], with. aggregated individual-level genotype and phenotype data from sequencing studies, including case-reports and large case series, published between 2001 and 2024. The database includes individual-level data for diagnoses across seven NDDs (ID, ASD, EP, CP, ADHD, schizophrenia, and bipolar disorder), and their corresponding single-gene causative variant (loss-of-function, missense, or single-gene deletion), with *de novo*, inherited, or unknown inheritance included. Clinical diagnoses were taken from the original reports.  We analyzed 8,973 probands with at least one early-onset neurodevelopmental disorder (ID, ASD, EP, or CP) and a causative variant in one of 263 high-confidence NDD genes with inheritance patterns consistent with known disease mechanisms (Supplementary Table 1). Genes and individual filters were applied to enrich for genes with strong evidence for NDD association (Supplementary Fig. 1). High-confidence genes were defined using Gene Curation Coalition (GenCC) classifications (accessed May 28, 2025) [2] and CP literature [3]. All 263 genes met GenCC criteria for Definitive or Strong evidence supporting their association with NDD related disease (Supplementary Table 1). Each gene was represented by at least ten individuals.

###### *Validation cohort*

The validation cohort consisted of 19,704 individuals who underwent clinical exome or genome sequencing at GeneDx with results reported between January 2015 and June 2025. This study was conducted under a GeneDx research protocol approved by the Western Institutional Review Board, Study Number 1169768, WIRB Pro Number 20162523, which permitted a waiver of consent. Eligible individuals included in this study had a confirmed likely pathogenic or pathogenic variant based on ACMG criteria [4] in at least one of the 263 genes of interest and at least one of 362 Human Phenotype Ontology (HPO) terms corresponding to the four NDDs (Supplementary Table 4). Diagnoses were obtained from clinician-submitted requisition forms and clinical notes and converted to Human Phenotype Ontology (HPO) terms by trained medical abstractors, as previously described [5]. Genes represented by fewer than 10 individuals were excluded. The final dataset included 234 genes represented by ≥10 individuals, with gene-level phenotype frequency profiles summarized in Supplementary Table 5.

#### Supplementary Fig.1

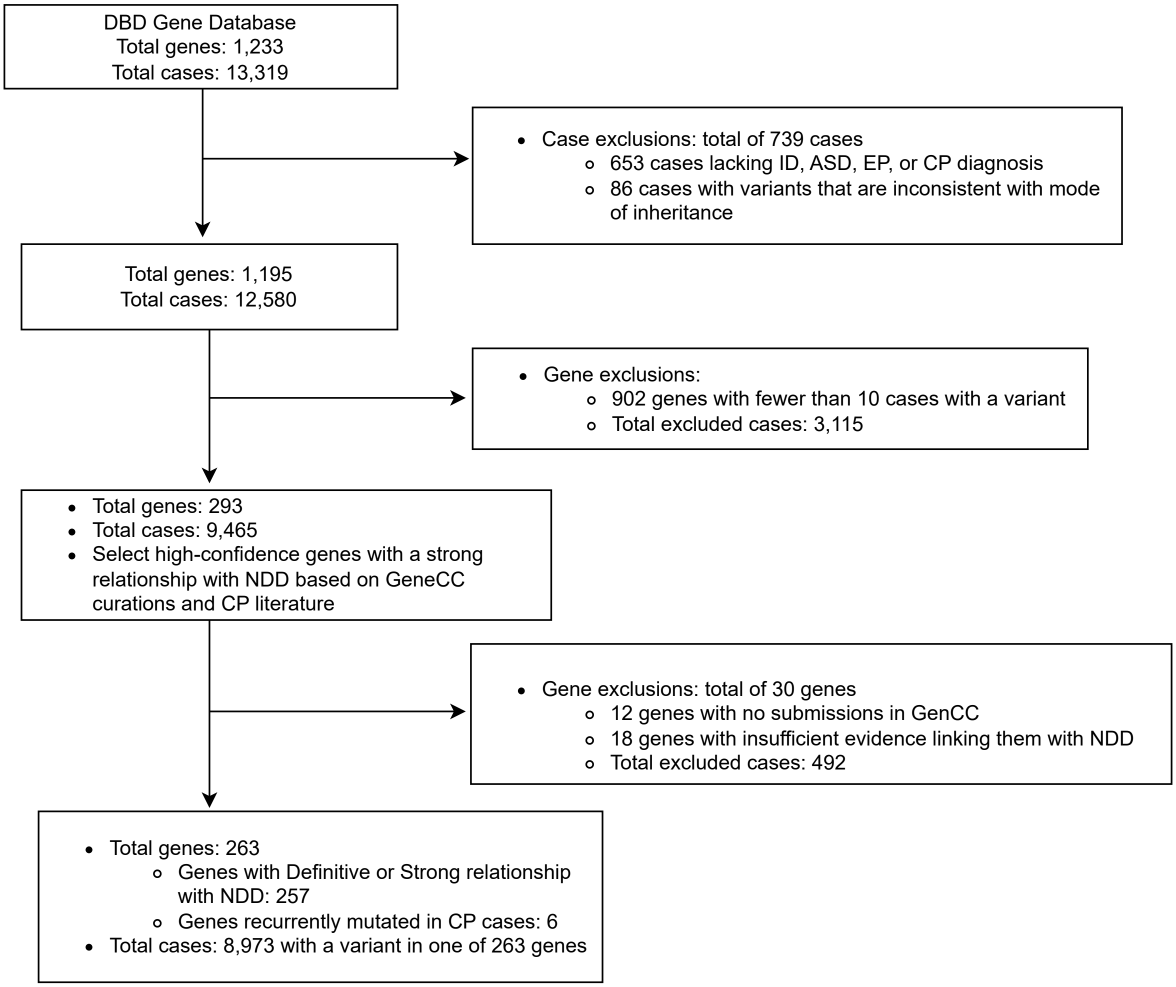


**Supplementary Fig.1 Selection of 263 high–confidence NDD genes and 8,973 individuals included in the discovery cohort from the DBD Gene Database.** High–confidence NDD genes were identified based on classifications curated by the Gene Curation Coalition (GenCC) [2] (accessed May 28, 2025) and supporting evidence from the CP literature [3].

#### Supplementary Fig.2

**A**

```
Cophenetic correlation for Euclidean distance and single linkage is 0.6913.
Cophenetic correlation for Euclidean distance and complete linkage is 0.7710.
Cophenetic correlation for Euclidean distance and average linkage is 0.7831.
Cophenetic correlation for Euclidean distance and weighted linkage is 0.7147.
Cophenetic correlation for Euclidean distance and ward linkage is 0.5913.
Cophenetic correlation for Chebyshev distance and single linkage is 0.6814.
Cophenetic correlation for Chebyshev distance and complete linkage is 0.6959.
Cophenetic correlation for Chebyshev distance and average linkage is 0.7794.
Cophenetic correlation for Chebyshev distance and weighted linkage is 0.6973.
Cophenetic correlation for Mahalanobis distance and single linkage is 0.5585.
Cophenetic correlation for Mahalanobis distance and complete linkage is 0.5534.
Cophenetic correlation for Mahalanobis distance and average linkage is 0.6979.
Cophenetic correlation for Mahalanobis distance and weighted linkage is 0.6835.
Cophenetic correlation for Cityblock distance and single linkage is 0.6873.
Cophenetic correlation for Cityblock distance and complete linkage is 0.7314.
Cophenetic correlation for Cityblock distance and average linkage is 0.7885.
Cophenetic correlation for Cityblock distance and weighted linkage is 0.7566.
****************************************************************************************************
Highest cophenetic correlation is 0.7885, which is obtained with Cityblock distance and average linkage.
```

**B**

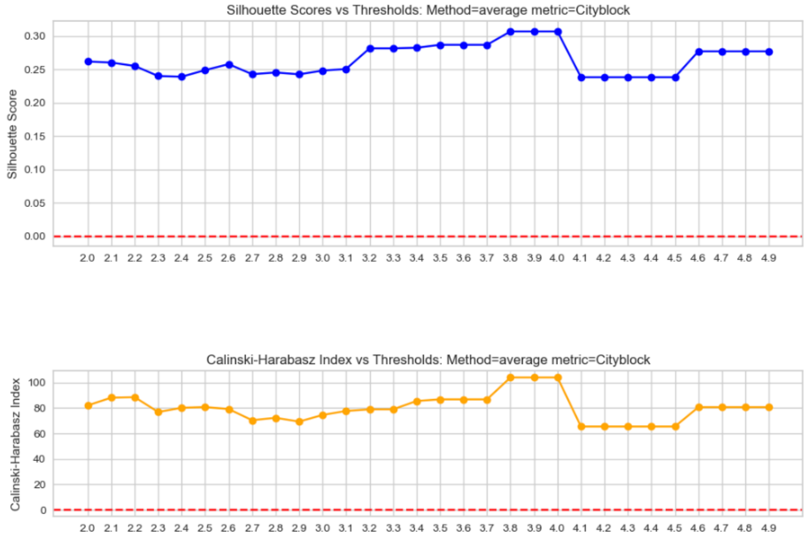


**Supplementary Fig.2 Optimal hierarchical clustering method selection. A** The optimal combination of distance metrics and linkage method for clustering was determined by performing hierarchical clustering on the data with various combinations, selecting the one that resulted in the highest cophenetic correlation coefficient. **B** After hierarchical clustering, flat clusters were generated by cutting the dendrogram at a chosen distance threshold, producing distinct, non–nested clusters. We selected the threshold that maximized two cluster–quality metrics–the Silhouette Score and the Calinski–Harabasz Index. Testing a range of linkage distances showed that thresholds between 3.8 and 4.0 produced the highest cluster quality, corresponding to peak values of both metrics. We selected a threshold of 4.0, which resulted in seven clusters (including a cluster with one gene).

#### Supplementary Fig.3

### A

ID cluster

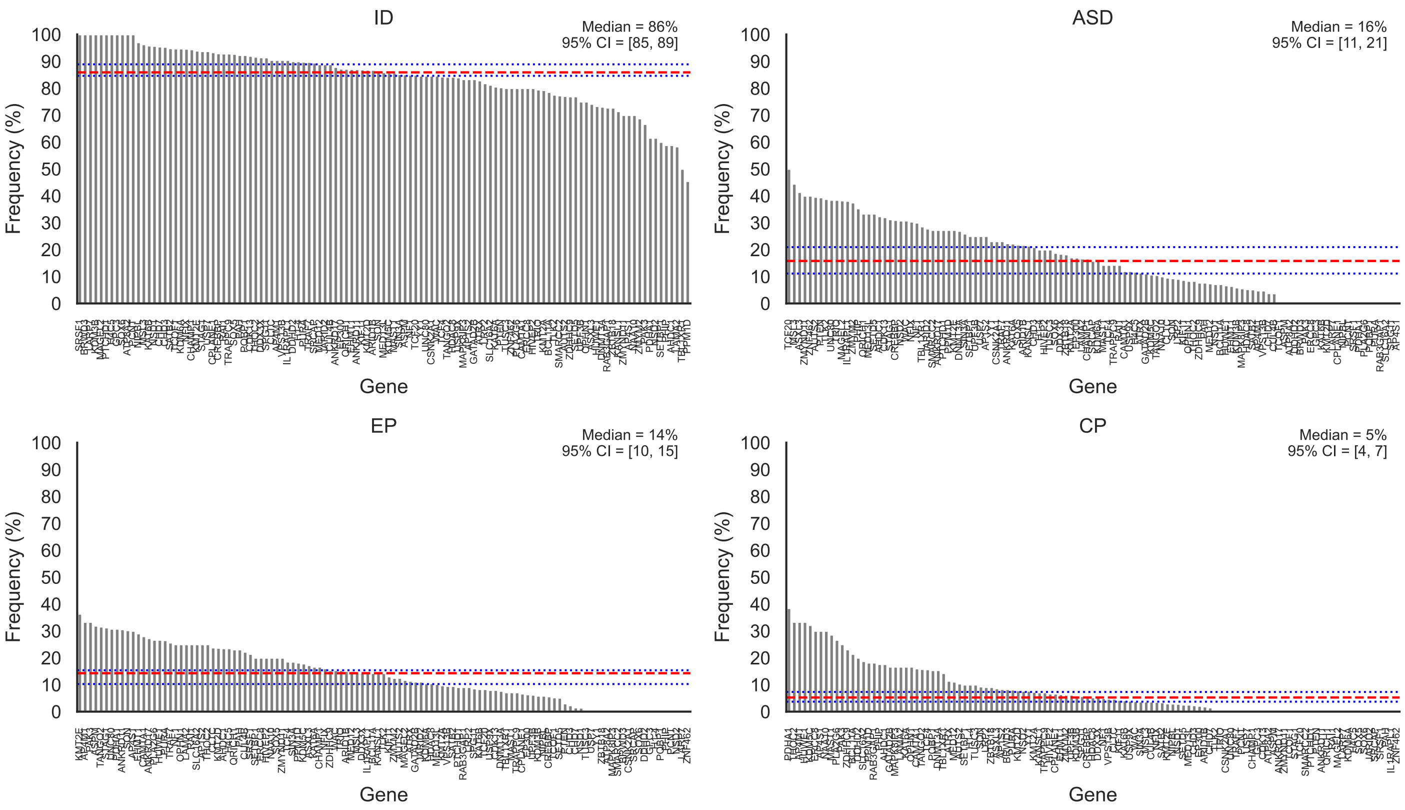

#### Supplementary Fig.3

ASD–ID cluster

**B**

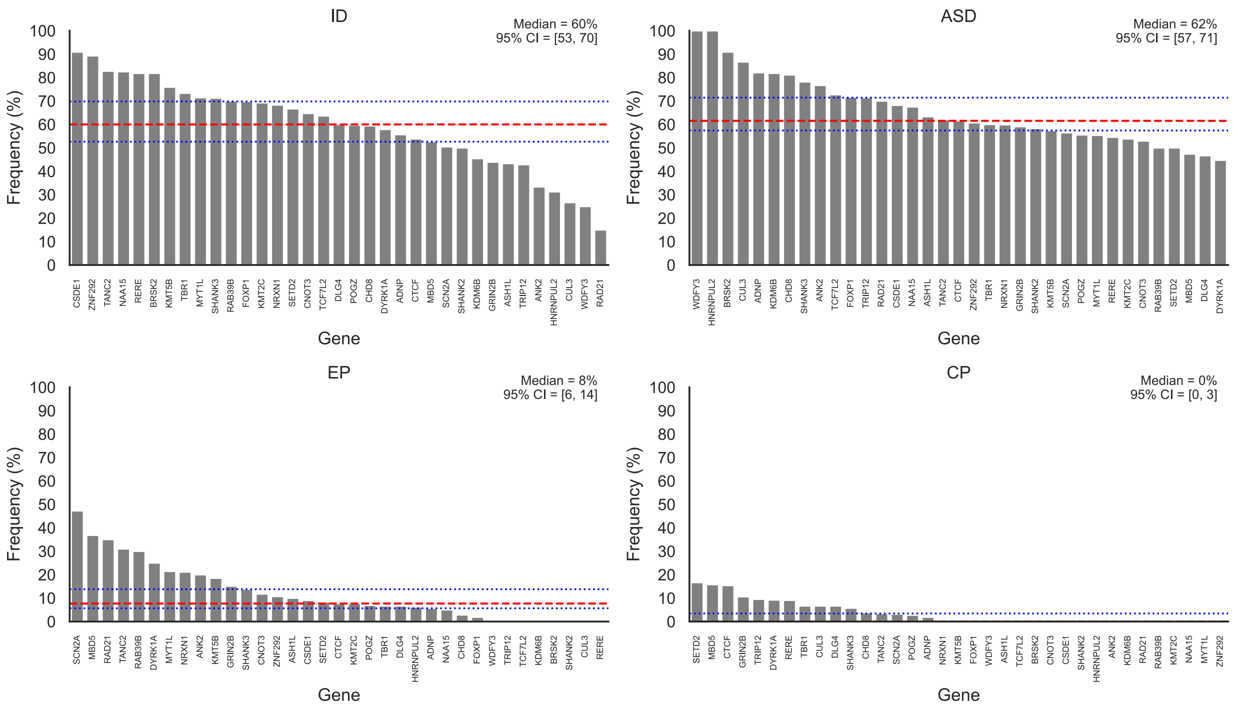

#### Supplementary Fig.3

**c**

ID–EP cluster

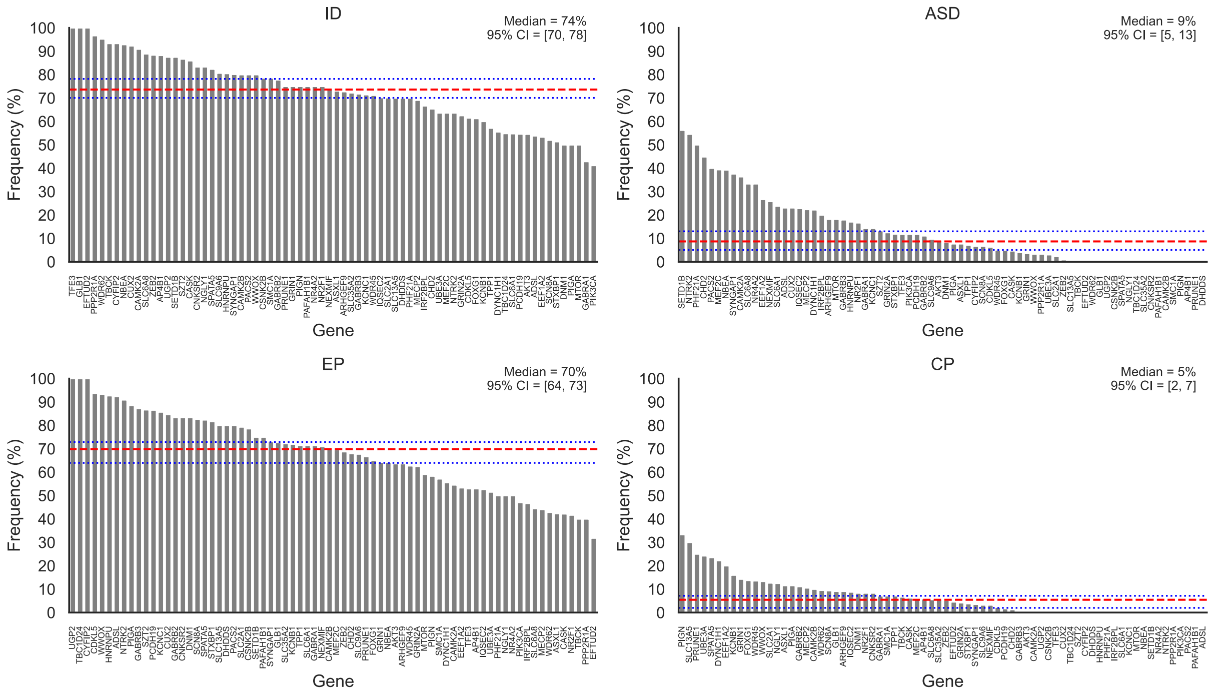

#### Supplementary Fig.3

**D**

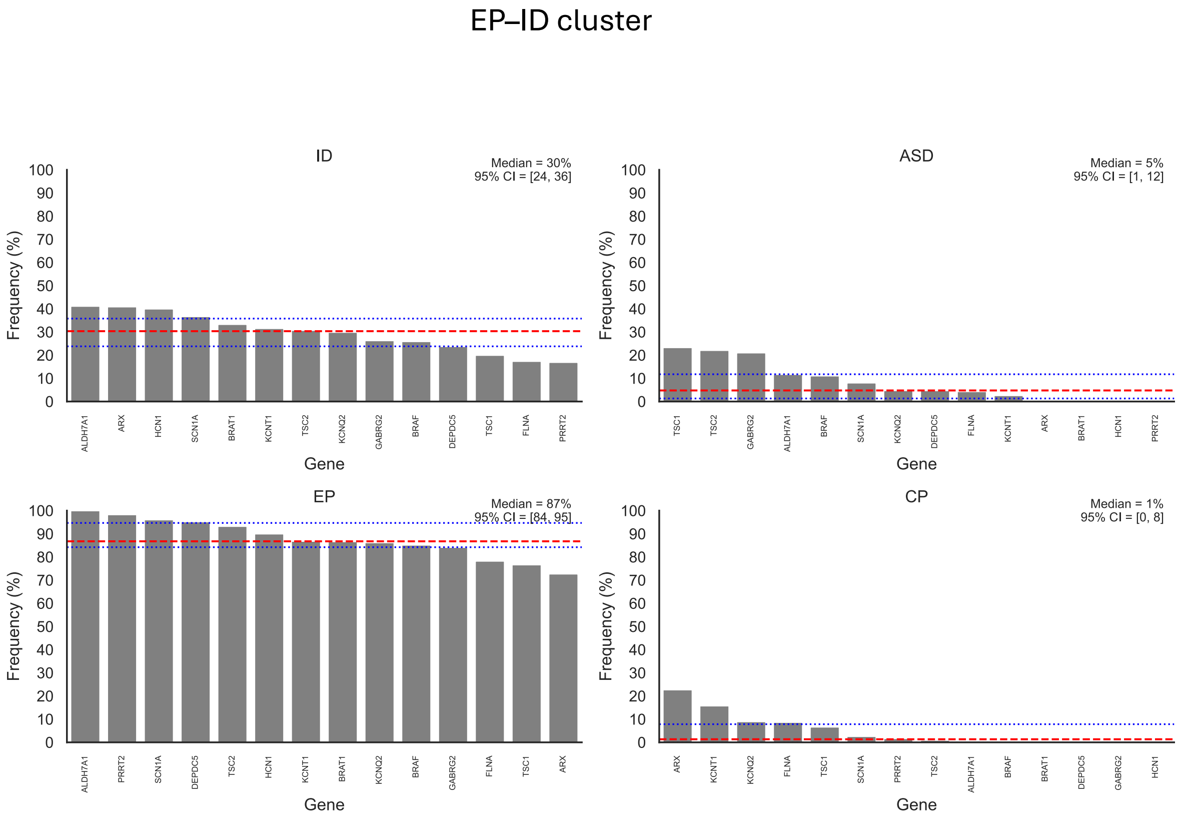

#### Supplementary Fig.3

**E**

ID–CP cluster

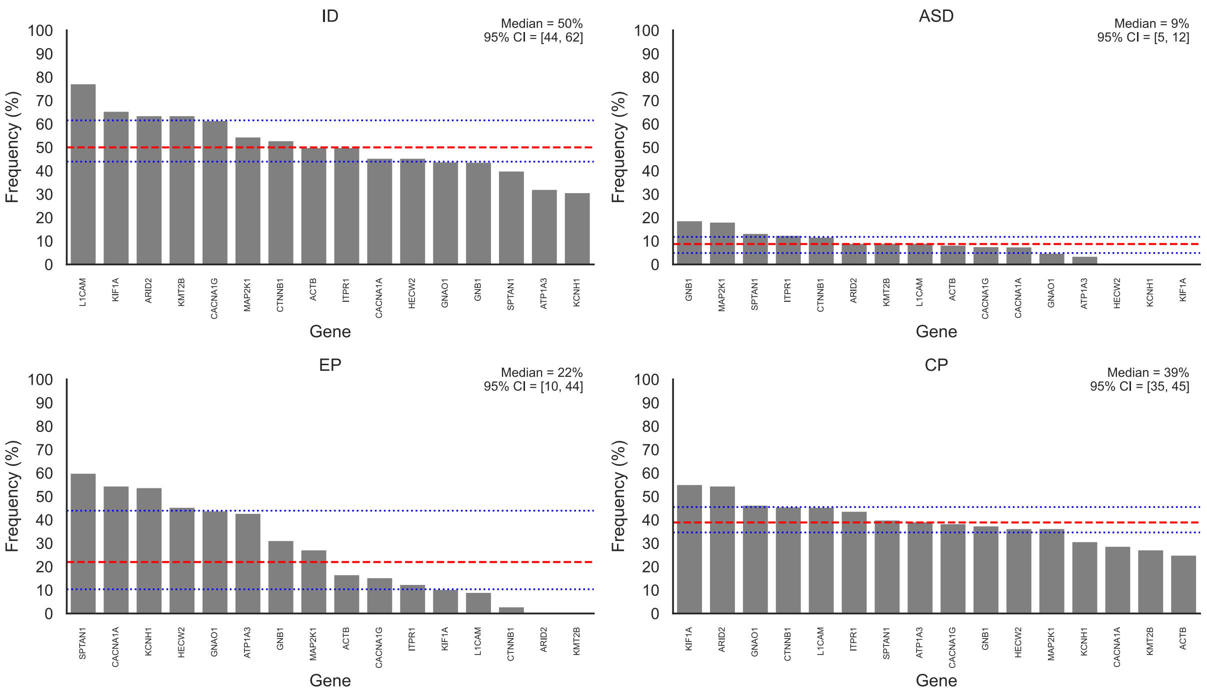

**Supplementary Fig.3**

**F**

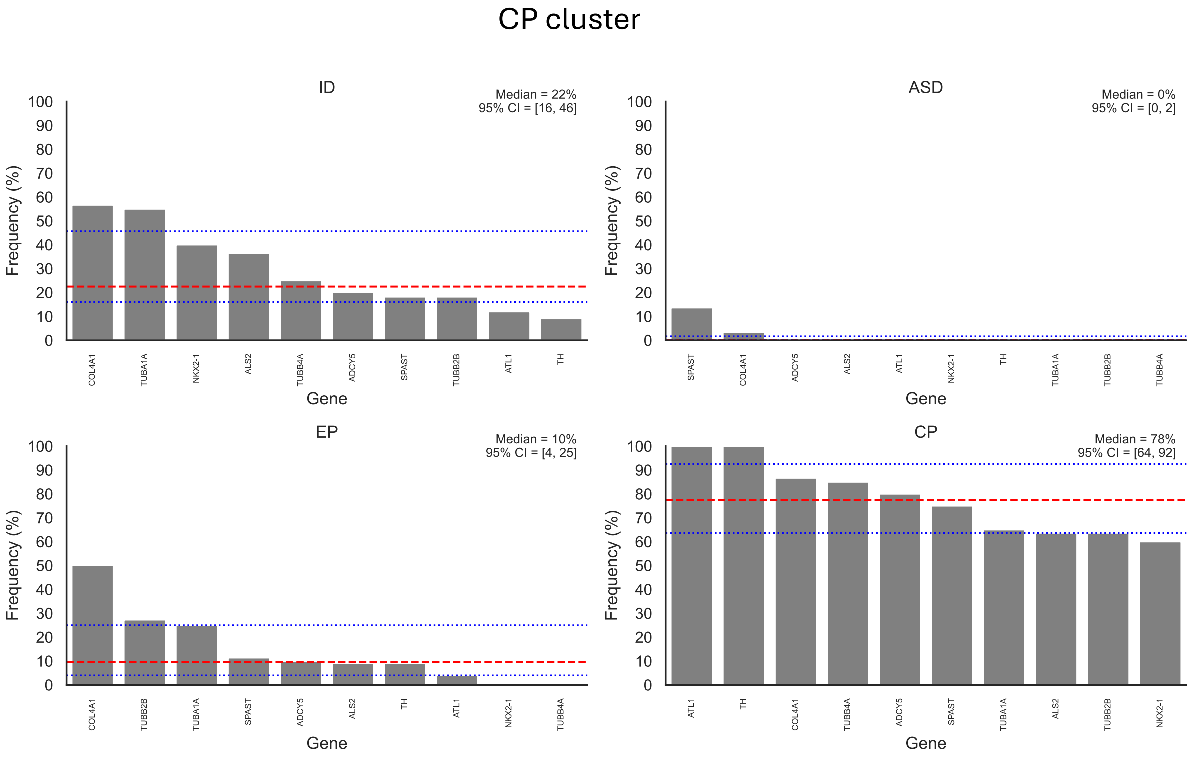


**Supplementary Fig.3 Frequency of NDD phenotypes across genes within each cluster in the discovery dataset.** The dotted red line indicates the median disorder frequency (%), and blue lines represents the 95% CI of disorder frequencies for genes in each cluster (A-F).

**Supplementary Fig.4**

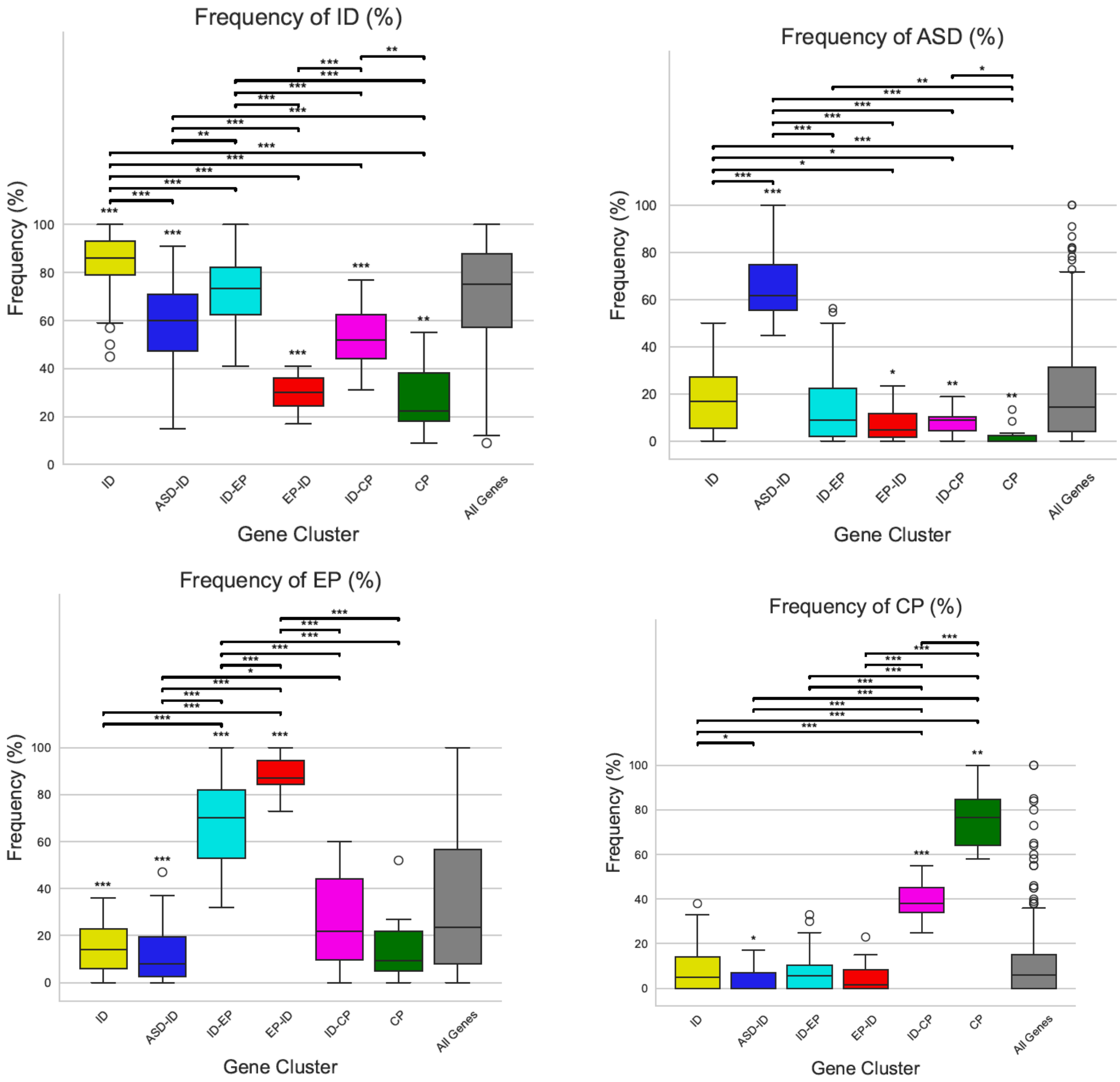


**Supplementary Fig.4 Disorder frequencies significantly differed between gene clusters.** Frequencies of each disorder were compared across gene clusters in the discovery dataset. Boxplots show the distribution of disorder frequencies for genes in each cluster across the four disorders (ID, ASD, EP, and CP). Statistical significance was assessed using two tests: (1) a one–sample Wilcoxon signed–rank test with Benjamini–Hochberg correction to compare the distribution of gene-level disorder frequencies within each cluster to the overall median frequency across all genes, with significant differences marked by asterisks above boxplots; and (2) Pairwise Mann–Whitney U test with to compare clusters, with significant results shown by connecting lines and asterisks. For each disorder, p values from both tests were jointly corrected for multiple testing using the Benjamini–Hochberg false discovery rate procedure. * $p < 0.05$, ** $p < 0.01$, *** $p < 0.001$.

#### Supplementary Fig. 5

**A**

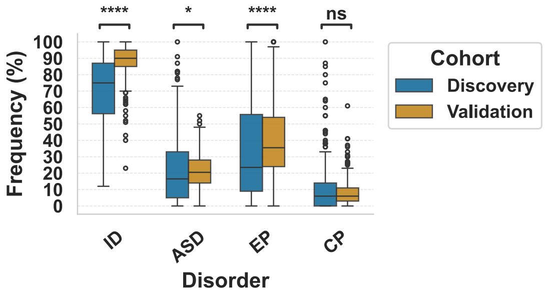


**B**

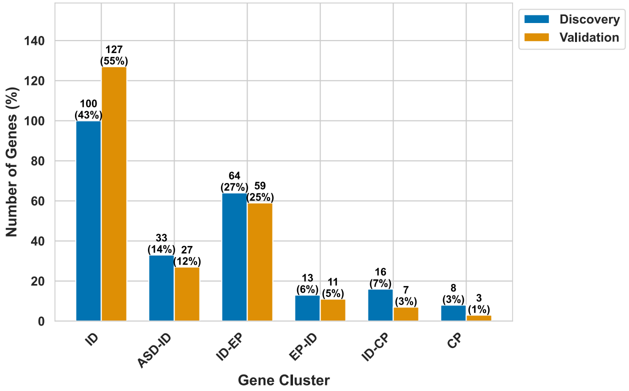


**C**

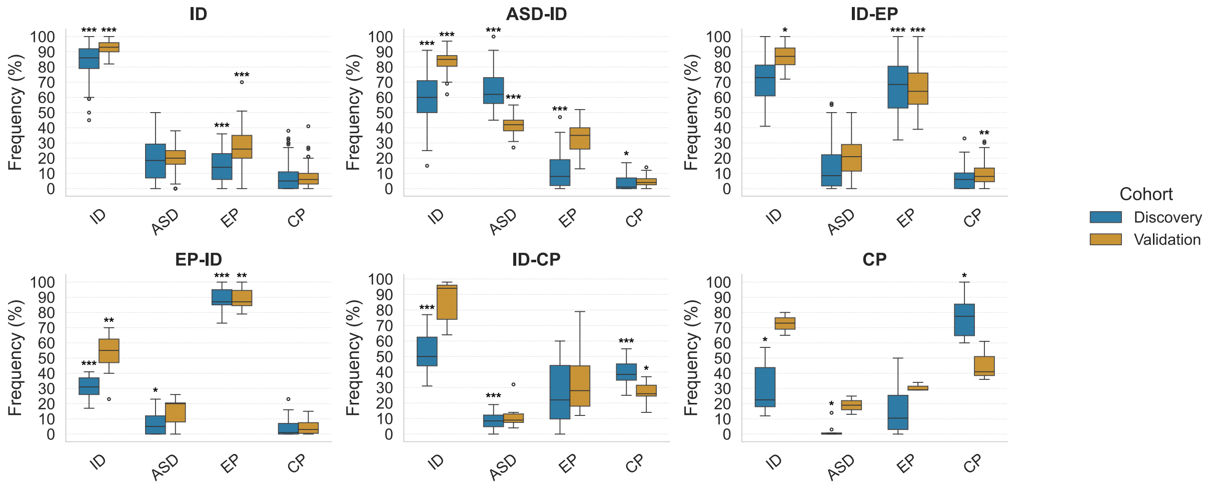


**Supplementary Fig. 5 Comparative distributions of disorder frequencies and gene cluster profiles across 234 common genes in the discovery and validation datasets. A** Boxplots show of disorder frequency distribution across 234 genes in the discovery and validation cohorts. Differences between cohorts were tested using the Mann–Whitney U test, with significant p–

values marked by asterisks (*). **B** Bar plots show the number of genes from the discovery (blue) and validation (orange) datasets assigned to each cluster. Similar proportions across clusters indicate consistent cluster structure between datasets. **C** Boxplots compare disorder frequencies by gene cluster for the 234 shared genes. Within each cohort, cluster-specific frequencies were tested against the overall gene set using a two-sided one-sample Wilcoxon signed-rank test. P-values were corrected for multiple testing across all cluster–disorder comparisons using the Benjamini–Hochberg procedure. Significant results are indicated by asterisk (* adjusted $p < 0.05$, ** adjusted $p < 0.01$, *** adjusted $p < 0.001$).

**Supplementary Fig. 6**

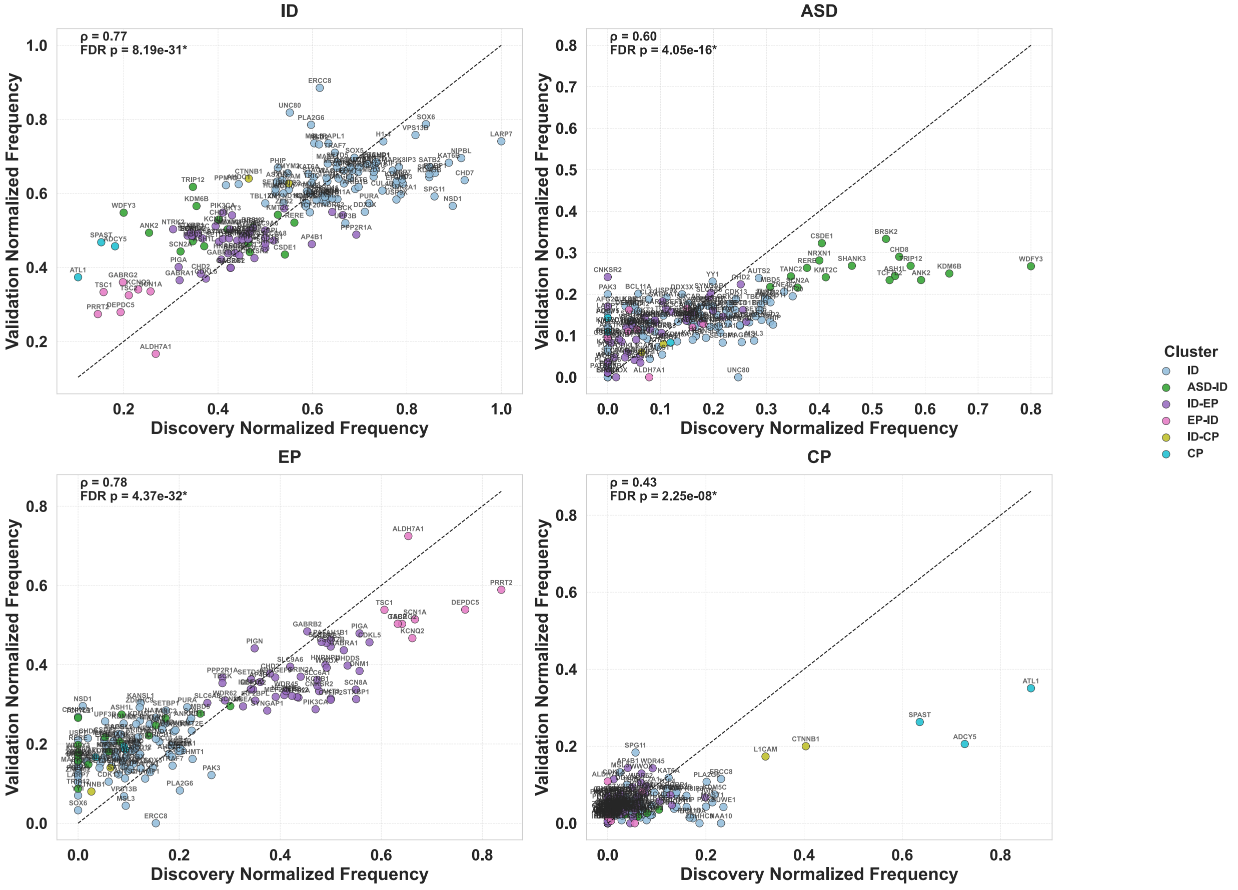


**Supplementary Fig. 6 Correlations of gene–level normalized disorder frequencies between discovery and validation datasets.** Scatter plots show correlations of normalized disorder frequencies for 155 genes with consistent cluster assignments in both discovery and validation datasets. The x–axis indicates normalized disorder frequencies from discovery, and the y–axis shows those from validation datasets. Each point represents a single gene, colored by cluster assignment. The dashed diagonal line denotes the identity line, where genes would fall if frequencies were identical between datasets. For each disorder, the Spearman correlation coefficient (ρ) and the FDR–corrected p–values are reported, with statistically significant correlations marked by an asterisk. These results highlight the concordance of gene–level disorder representation across the two cohorts.

#### Supplementary Fig. 7

**A**

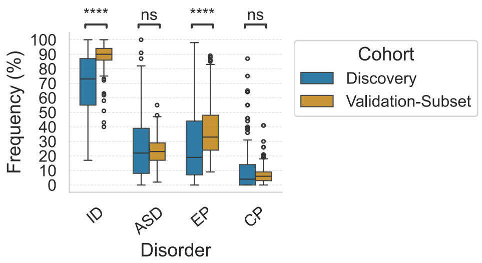


**B**

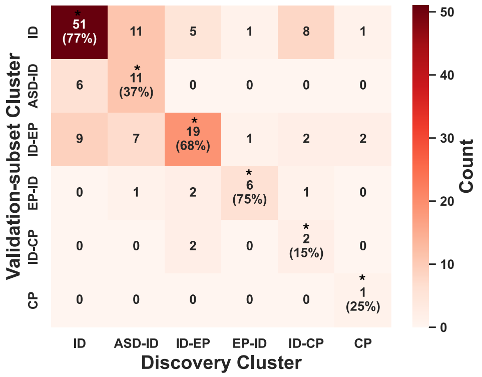


**C**

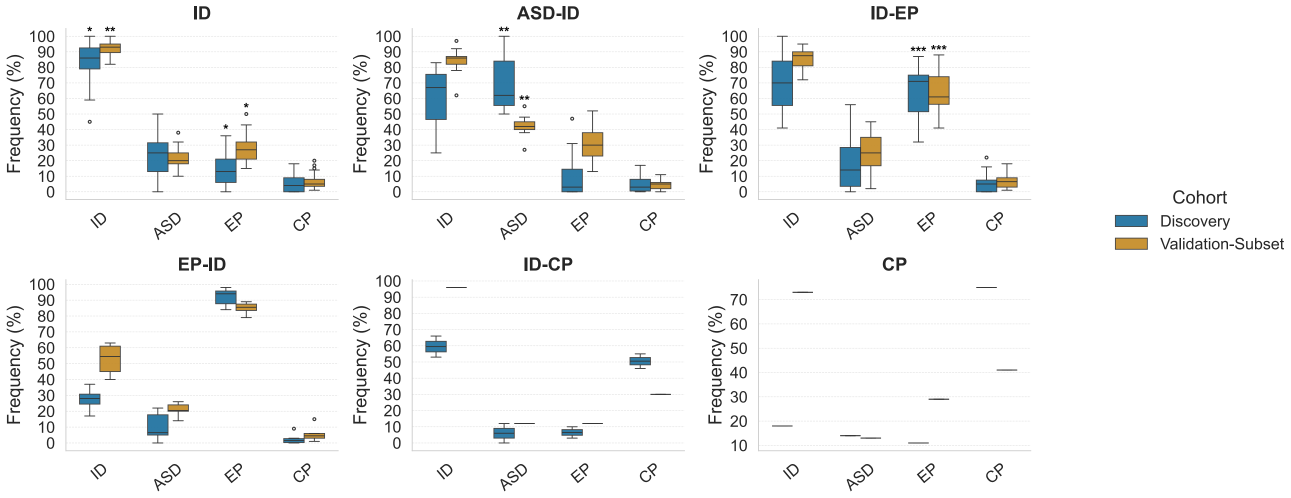


**Supplementary Fig.7 Validation of gene clusters using the validation data subset. A** Boxplots show disorder frequency distributions for 149 genes in the discovery and validation subsets. Significant cohort differences for each disorder are indicated by asterisks. **B** Confusion matrix showing cluster assignments based on disorder frequencies in the validation cohort subset.

Diagonal elements indicate genes assigned to the same cluster in both datasets (90/149 genes; 60%, permutation test, $p < 0.0001$), with per-cluster percentages in parentheses. Clusters with more concordant assignment than expected by chance are indicated with asterisks (adjusted permutation $p < 0.05$). Off–diagonal elements represent genes assigned to different clusters between the two datasets. **C** Boxplots compare disorder frequencies across clusters for the 90 genes with concordat cluster labels. Significance reflects deviation from the cohort-wide median based on two-sided Wilcoxon signed-rank tests with Benjamini–Hochberg correction (*: $p < 0.05$; **: $p < 0.01$, ***: $p < 0.001$). No p-values were computed for the CP cluster due to single concordant gene.

**Supplementary Fig.8**

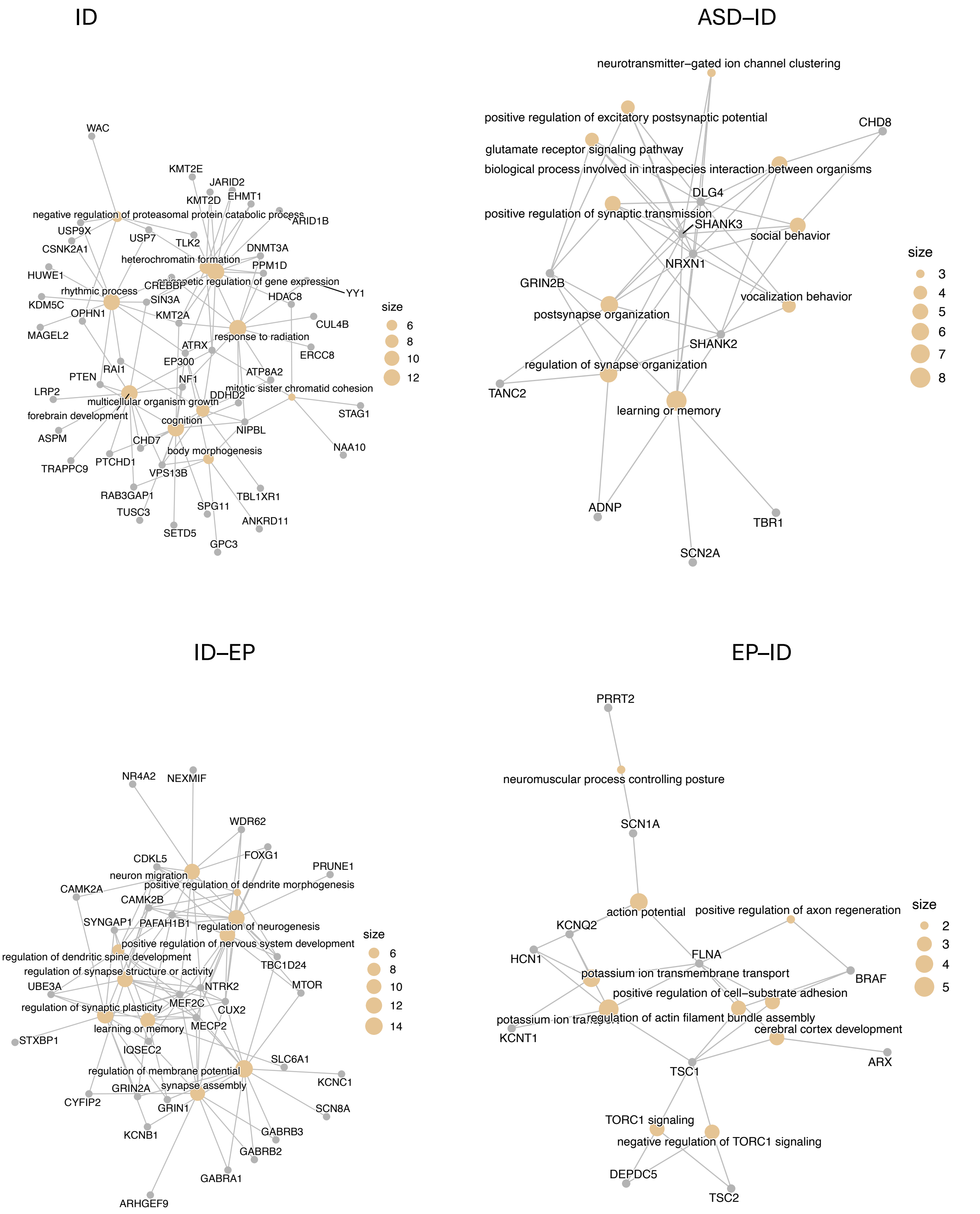

**Supplementary Fig.8**

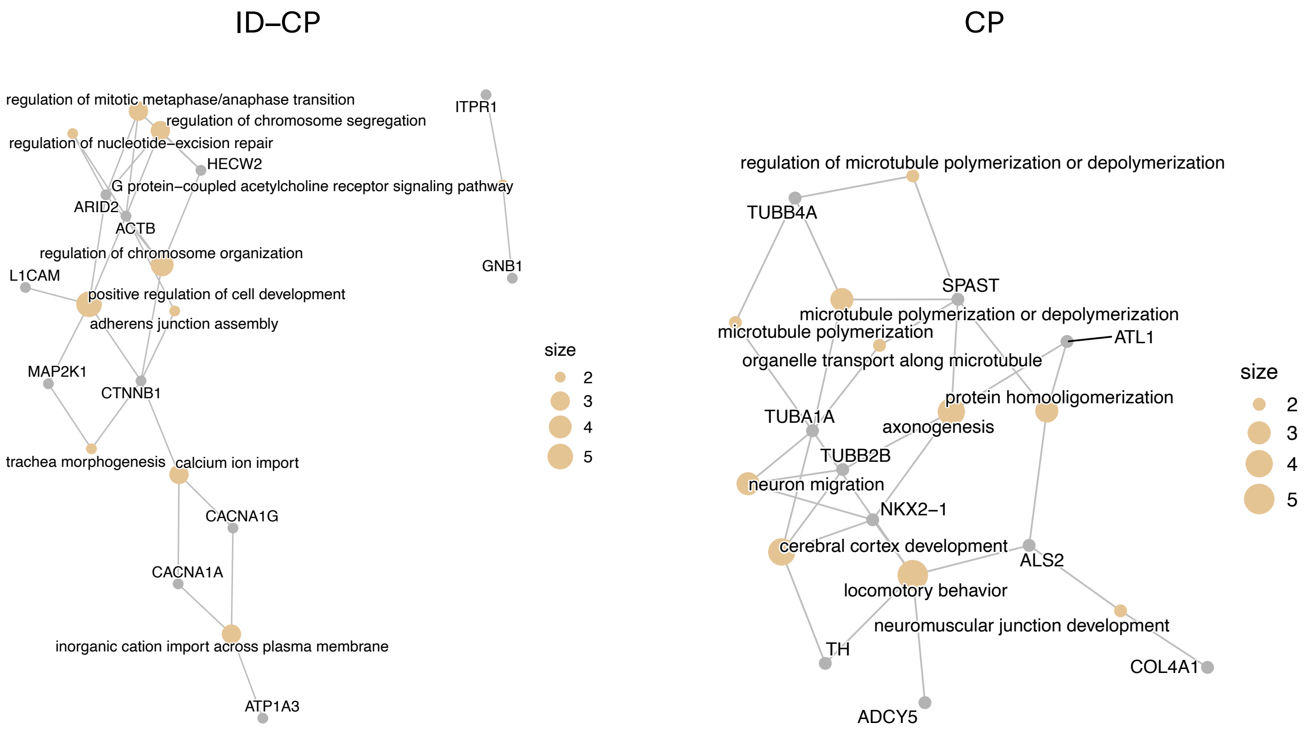


**Supplementary Fig.8 Top 10 enriched GO biological processes are shared by multiple genes.** Genes linked to enrichment of the top 10 biological processes are plotted. The size of each node represents the number of genes by which the term is enriched; larger nodes indicate processes that are more prevalent in the gene set for the given cluster.

#### Supplementary Fig.9

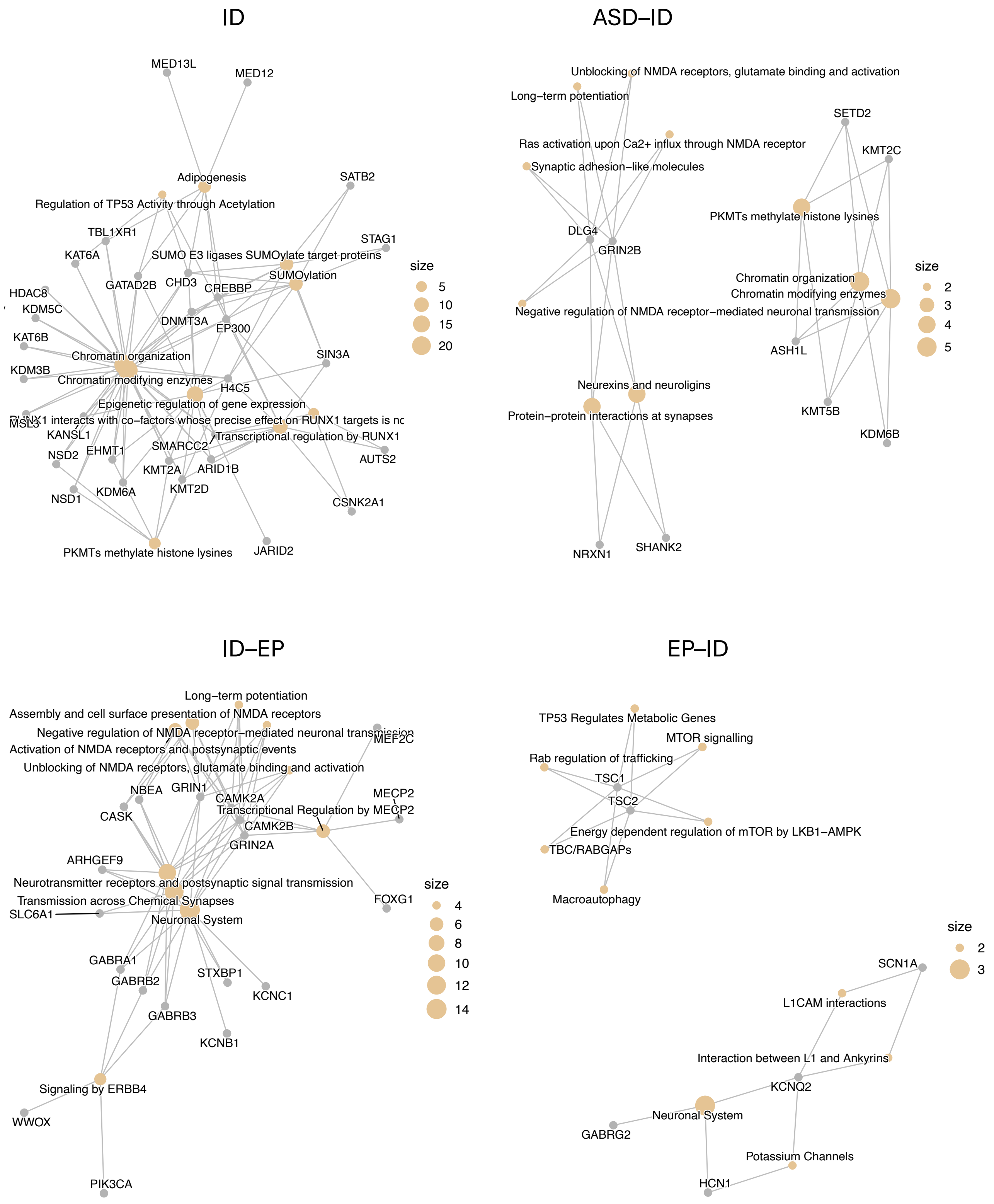

**Supplementary Fig.9**

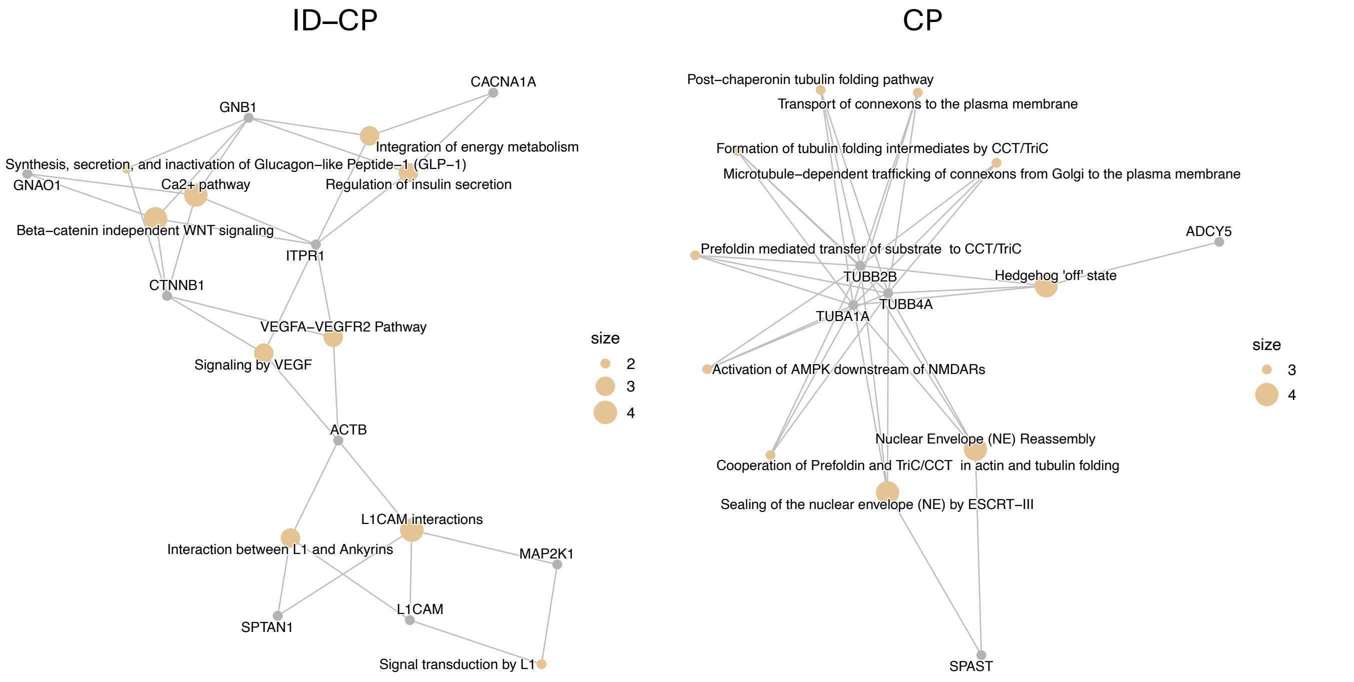


**Supplementary Fig.9 Top 10 enriched pathways are shared by multiple genes.** Genes linked to enrichment of the top 10 pathways are plotted. The size of each node represents the number of genes by which the term is enriched; larger nodes indicate pathways that are more prevalent in the gene set for the given cluster.

#### Supplementary Fig.10

**A**

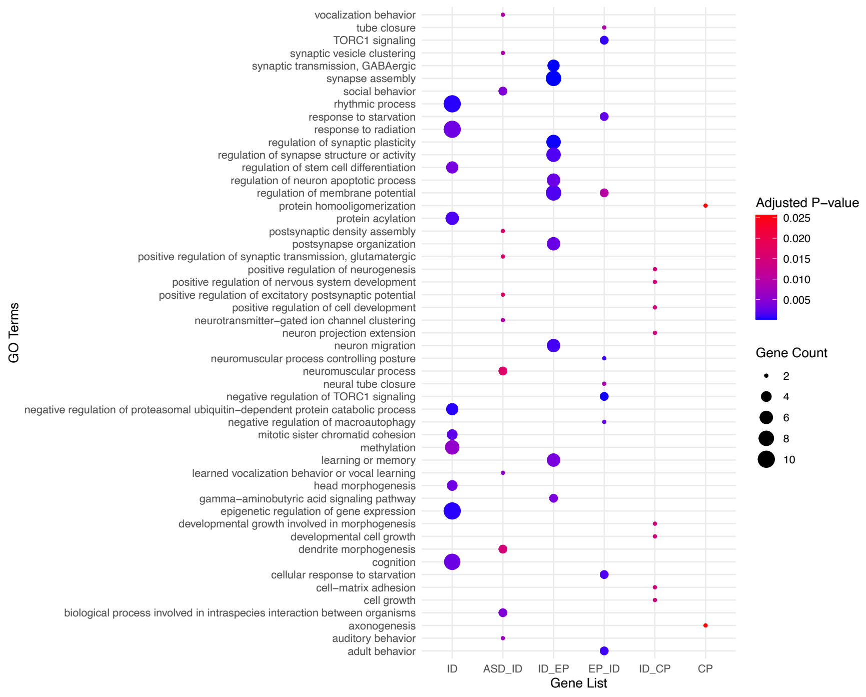


**B**

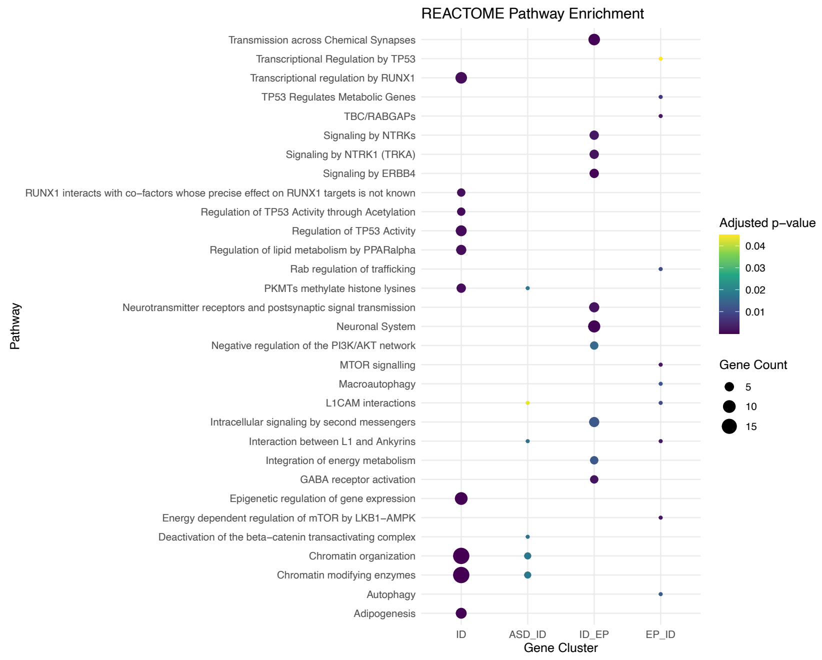


**Supplementary Fig.10 GO biological processes and pathways enriched within gene clusters comprising the 155 genes with consistent cluster assignments across the discovery and validation datasets. a,** The top 10 enriched biological processes with adjusted p–value < 0.05 are plotted. **b,** The top 10 significantly enriched Reactome pathways colored by adjusted p–values (<0.05). Gene clusters are shown on the x–axis. The size of each dot corresponds to the number genes in a pathway. No pathways were significantly enriched in the CP–ID or CP clusters.

**REFERENCES**


1. Gonzalez-Mantilla AJ, Moreno-De-Luca A, Ledbetter DH, Martin CL. A Cross-Disorder Method to Identify Novel Candidate Genes for Developmental Brain Disorders. JAMA Psychiatry. 2016;73:275–283.
2. DiStefano MT, Goehringer S, Babb L, Alkuraya FS, Amberger J, Amin M, et al. The Gene Curation Coalition: A global effort to harmonize gene-disease evidence resources. Genet Med. 2022;24:1732–1742.
3. van Eyk CL, Fahey MC, Gecz J. Redefining cerebral palsies as a diverse group of neurodevelopmental disorders with genetic aetiology. Nat Rev Neurol. 2023;19:542–555.
4. Richards S, Aziz N, Bale S, Bick D, Das S, Gastier-Foster J, et al. Standards and guidelines for the interpretation of sequence variants: a joint consensus recommendation of the American College of Medical Genetics and Genomics and the Association for Molecular Pathology. Genet Med. 2015;17:405–424.
5. Retterer K, Juusola J, Cho MT, Vitazka P, Millan F, Gibellini F, et al. Clinical application of whole-exome sequencing across clinical indications. Genet Med. 2016;18:696–704.
